# Antibody signatures against viruses and microbiome reflect past and chronic exposures and associate with aging and inflammation

**DOI:** 10.1101/2024.01.12.24301197

**Authors:** Sergio Andreu-Sánchez, Aida Ripoll-Cladellas, Anna Culinscaia, Ozlem Bulut, Arno R. Bourgonje, Mihai G. Netea, Peter Lansdorp, Geraldine Aubert, Marc Jan Bonder, Lude Franke, Thomas Vogl, Monique G.P. van der Wijst, Marta Melé, Debbie Van Baarle, Jingyuan Fu, Alexandra Zhernakova

**Author notes:** Correspondence to: Alexandra Zhernakova, Jingyuan Fu.

## Abstract

Prior encounters with pathogens and other molecules can imprint long-lasting effects on our immune system, potentially influencing future physiological outcomes. However, given the wide range of pathogens and commensal microbes to which humans are exposed, their collective impact on the health and aging processes in the general population is still not fully understood. In this study, we aimed to explore relations between exposures, including to pathogens, microbiome and common allergens, and biological aging and inflammation. We capitalized on an extensive repository of the antibody-binding repertoire against 2,815 microbial, viral, and environmental peptides in a deeply-phenotyped population cohort of 1,443 participants. Utilizing antibody-binding as a proxy for past exposures, we investigated their impact on biological aging markers, immune cell composition and systemic inflammation. This identified that immune response against cytomegalovirus (CMV), rhinovirus and specific gut bacterial species influences the telomere length of different immune cell types. Using blood single-cell RNA-seq measurements, we identified a large effect of CMV infection on the transcriptional landscape of specific immune cells, in particular subpopulations of CD8 and CD4 T-cells. Our work provides a broad examination of the role of past and chronic exposures in biological aging and inflammation, highlighting a role for chronic infections (CMV and Epstein-Barr Virus) and common pathogens (rhinoviruses and adenovirus C).

**Highlights:**

- The study provides a broad association of antibody reactivity with biomarkers of aging and inflammation
- It shows that anti-CMV, rhinovirus and gut antimicrobial antibody reactivity relate to telomere length
- CMV infection associates to the telomere length of CD45RA+CD57+ cells in a sex-dependent manner
- CMV influences the transcriptomic landscape of CD8+ T effector memory and cytotoxic CD4+ cell populations
- Anti-Epstein-Barr-Virus and anti-adenoviral responses are associated with higher circulating IL-18BP concentrations

## Introduction

The dramatic increase in human lifespan over the last century has not been accompanied by a similarly rapid increase in health span, leading to a large social and economic burden due to the susceptibility of older people to disease. Aging is associated with an increased incidence of chronic conditions, such as cardiovascular diseases, cancer, diabetes, and neurodegenerative disorders (1,2). Immunosenescence—which represents a series of detrimental changes in the immune system (3)—is an accelerator of age-related diseases, with known hallmarks of reduced response to new antigens, accumulation of memory T-cells, and low-grade systemic inflammation that is termed “inflammaging” (4).

The complex interplay between exposures and the human immune system plays a critical role in determining cellular homeostasis throughout human life. Both genetic and environmental factors influence the behavior of the immune system via molecular recognition and epigenetic imprinting (5). A wide range of exposures, including to environmental factors (e.g. pollen, pollution, and interaction with animals), lifestyle factors (e.g. smoking and diet), and microorganisms such as bacteria, fungi, and viruses, influence the function of the immune system and can have long-term effects on an individual’s health. Previous studies have emphasized the role of latent infections with herpesviruses in immunosenescence via establishment of a life-long persistent infection in the host that may reactivate under certain conditions (6). Common herpesviruses infections include herpes simplex viruses, Epstein-Barr Virus (EBV) and human cytomegalovirus (CMV). The significance of these viruses for human health is further highlighted by work showing that presence of EBV antibodies elevates the risk of developing multiple sclerosis, an autoimmune condition affecting the nervous system (7), and by a link between herpes zoster infection and development of Alzheimer’s disease (8). In addition, CMV has been associated to the immune risk profile in the elderly, with CMV associated with an inverted CD8/CD4 ratio, a hallmark of immunosenescence (9).

The humoral immune system, which includes antibodies selected to recognize a specific antigen, comprises memory B-cells that continue to produce antibodies even after an immune reaction has ended. Analyzing an individual’s total antibody repertoire can therefore provide insights into the past and current exposures that have triggered an immune response. This study aimed to investigate how such microbial exposures modify immune homeostasis and lead to aging-like features, such as becoming more proinflammatory, and how this is connected with markers of biological aging. To do so, we leveraged antibody epitope repertoires in the Dutch population cohort Lifelines-DEEP (LLD) (10), that were profiled using two previously characterized Phage-display Immunoprecipitation Sequencing (PhIP-Seq) libraries (11–13). Using large-scale omics datasets available for the same cohort (n=1,443), including cytokine concentrations, bulk- and single-cell transcriptomics, DNA methylation, blood metabolomics (15) and aging biomarkers, such as signal joint T-cell receptor excision circles (sj-TRECS) and telomere lengths (TLs) from blood immune cell types (16), we explored the relationships between past exposures and biological aging and systemic inflammation biomarkers. This identified multiple downstream effects of past infections on biological aging, systemic immune responses, and the transcriptional activity of specific immune populations.

## Results

### TLs of immune cell types associate with specific antibody-bound peptides

To study previous and current exposures to commensal and pathogenic bacteria and viruses we assessed the reactivity of serum antibodies against two previously developed libraries of phage-displayed peptide antigens. In a previous study in the Dutch LLD cohort (13) PhIP-Seq was used to profile two antigen libraries. These libraries consisted of peptides derived from gut microbiota (both commensal species and common pathogens) (11); epitopes from common dietary allergens, phages and viruses; and all B-cell antigens deposited in the IEDB (immune epitope database) (12). Extensive IgG antibody responses against both libraries were observed in the study population (n=1,443) (13), and we used the 2,815 responses frequently shared among participants (detectable in 5–95% of individuals) for further analysis (13) [**Supplementary Table 1**].

To investigate the relationship between previous and current exposures (which could not be separated in this data) and aging, we explored various subsets of participants using different available biomarker information [**Figure 1A**]. We explored four types of aging biomarkers: TLs of six different blood cell types were measured by Flow-FISH: granulocytes, lymphocytes, B-cells (CD45RA+CD20+), naïve T-cells (CD45RA+CD20−), memory T-cells (CD45RA−) and NK-cells/fully differentiated T-cells (CD45RA+CD57+). DNA methylation age estimated using Hannum (17), Weidner (18) and Horvath (19) methods. Metabolic age, calculated from the blood NMR metabolomics profile using two previously published methods: MetaboAge, which predicts chronological age (20) and MetaboHealth, which predicts all-cause mortality risk (21). Thymic T-cell maturation function, which was measured by the expression level of its by-products, sj-TRECS. The sample overlap for the different biomarkers is displayed in [**Figure 1B**].

**Figure 1.**
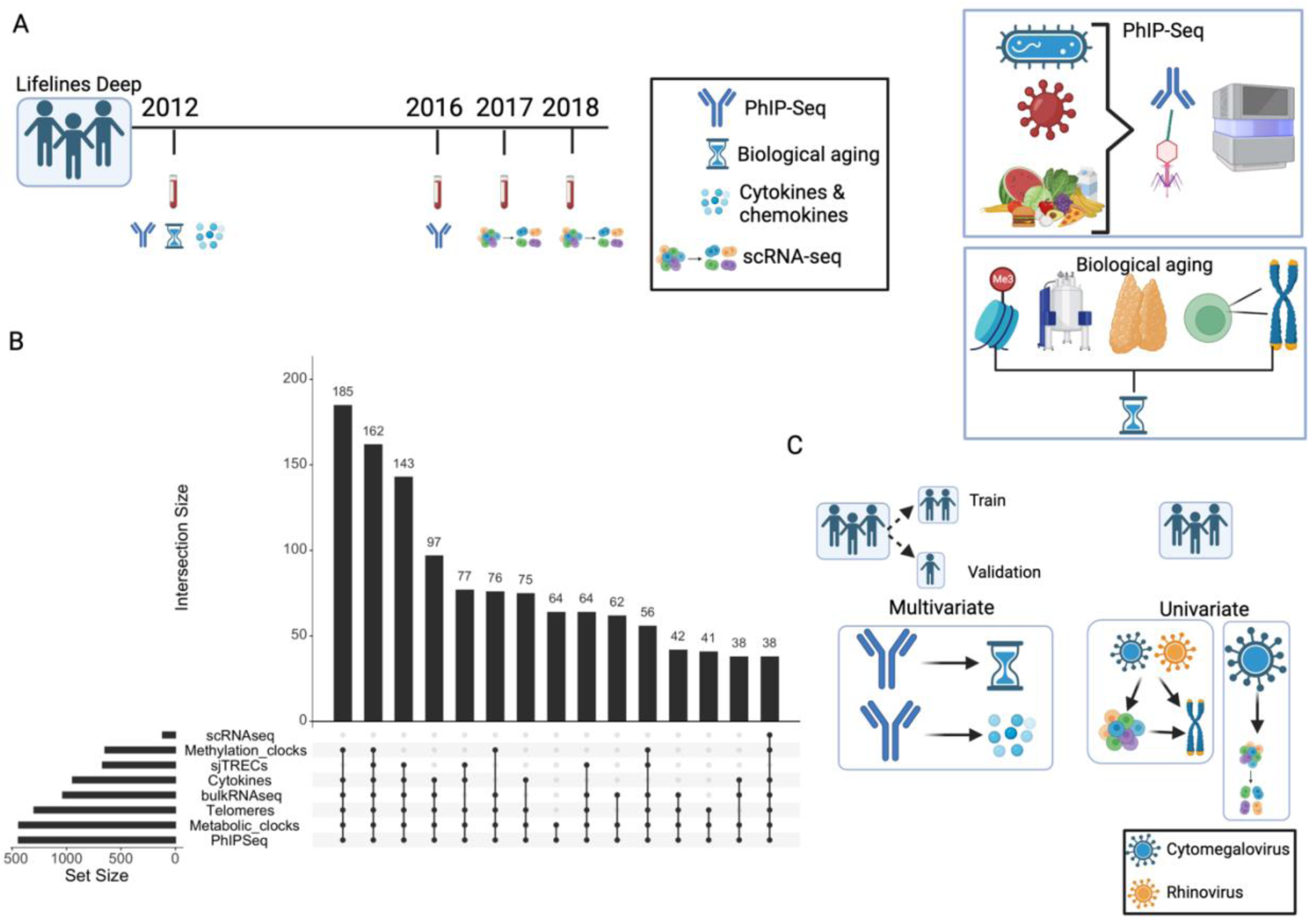
Data and methodological framework. **A.** Data from 1,443 participants of the Dutch cohort Lifelines-DEEP were used to explore the relationship of past and chronic exposures, inflammation and aging. PhIP-Seq was used to profile immune responses against 2,815 peptides. Fourteen circulating cytokines were available for 939 participants for whom we had PhIP-Seq data. Telomere lengths from six blood cell types (n=1,243), biological aging clocks (methylation clock (n=641), metabolomic clock (n=1,437)) and sj-TRECS (n=633) were used to investigate aging. Bulk blood (n=1,173) and peripheral blood mononuclear cell (PBMC) single-cell gene expression (n=119) were also obtained. **B.** Upset plot of common data subsets showing the number of samples with overlapping data layers. **C.** Analysis framework. Participants were split into a training and replication set. Univariate and multivariate techniques were applied to relate exposures (PhIP-Seq) to the other omics-derived aging and immunological biomarkers. Using the whole population, we explored the relationship of cytomegalovirus and rhinoviruses with cell counts and telomere length. Cytomegalovirus, in particular, was related to single-cell gene expression changes measured in samples obtained 1–6 years after collection of blood for PhIP-Seq measurements.

In our previous study (13), we uncovered the widespread association of chronological age with antibody responses, with prevalence of 13% of antibody-bound peptides associated to chronological age. Among these peptides, pathogens showed a wide diversity of age-associated immune responses, including more herpesvirus prevalence with age: CMV (61 associated antibody-bound peptides), EBV (46 associations) or herpesvirus 1 (10 associations). By contrast, other pathogens such as *Staphylococcus aureus* (22 peptides), *Streptococcus pneumoniae* (14 peptides) and Influenza A (9 peptides) showed a decrease of prevalence with age. To assess the correlation between antibody repertoires and biomarkers of biological aging (including TLs, metabolic age, methylation age, and sjTRECs) [**Figure 1B**], we first removed the variability attributed to chronological aging (see **Methods**) and then employed a hierarchical approach, ‘High-sensitivity pattern discovery in large, paired multi-omic datasets’ (HAllA) (22), that allows for the identification of blocks of highly correlated features, while maintaining a low false discovery. We ran HAllA on a ‘training’ set of the data, that encompassed 80% of the samples and then validated the significant associations on a separate ‘validation’ test-set of the remaining 20% of samples, applying partial correlations [**Figure 1C**].

We observed many negative associations between TL and antibody-bound peptides from CMV, gut microbiome, bacteriophages, virulence factors, EBV, an antigen from severe acute respiratory syndrome (SARS) (present at low frequency in the population, ∼5%), and the probiotic strain *B. breve* (UCC2003) (complete table in **Sup. Table 2.1**). Many of these associations (61/84) were further observed in the validation set (**Sup. Table 2.2**). Of these negative associations, the strongest were between 66 (out of a total of 77 peptides) antibody-bound peptides from CMV and shorter TLs in NK-cells/fully differentiated T-cells and lymphocytes [Figure 2A]. CMV is a common pathogen present in approximately 50% of the Dutch population (23), and is known to cause chronic unresolved infections. CMV infection was previously linked to substantial changes in blood cell composition and is considered a hallmark of immunosenescence (24).

**Figure 2.**
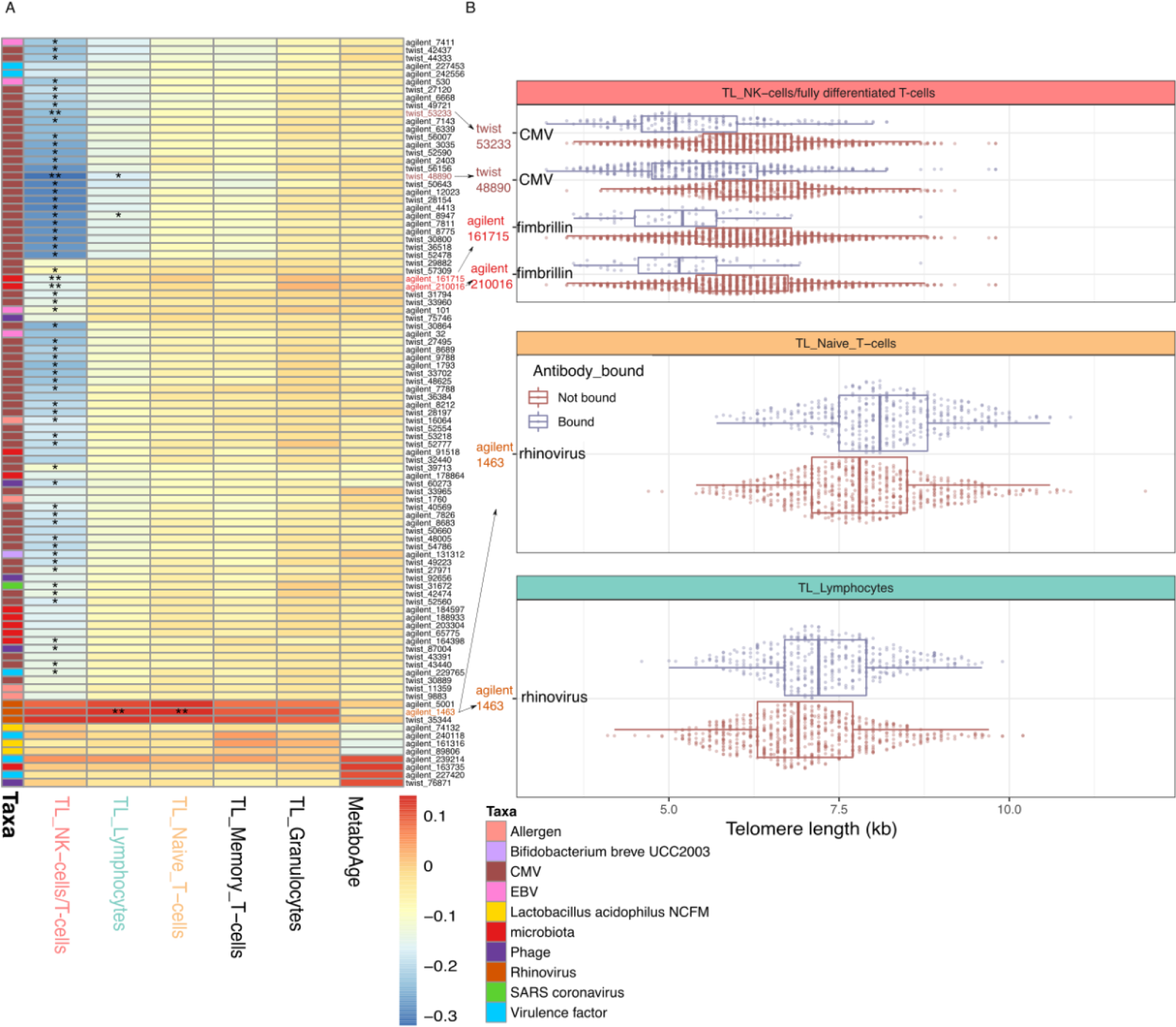
Associations between antibody-bound peptides and aging biomarkers. **A.** Heatmap showing correlation coefficients in the training dataset. Each row represents a peptide (colored according to taxonomic origin). Each column represents the telomere length (TL) of an immune cell type. * indicates significance (P < 0.05) in both the testing and training datasets when controlling for age and sex using partial correlation. ** indicates significance (P < 0.05) in both the training and validation datasets with additional control for CMV prediction (see text). **B.** Boxplots display the associations that remained significant in both the training and validation datasets after accounting for CMV serostatus. X-axis displays TL measurements determined using Flow-FISH for different cell populations. Y-axis indicates the peptides where the association was found. Color indicates whether the PhIP-Seq results predicted antibody-binding against the peptide.

In contrast to these negative associations, antibody-bound peptides from rhinovirus were positively correlated with the TLs of lymphocytes, naïve T-cells and NK-cells/fully differentiated T-cells. Rhinoviruses are respiratory pathogens that are usually responsible for causing the common cold.

In the training set, we further identified negative associations between antibody-binding against peptides and metabolic age. This included peptides belonging to the probiotic strain *Lactobacillus acidophilus* and to *Streptococcus*. In addition, we found associations of peptides from virulence factors from *Coprococcus* and *Mycoplasma pneumoniae* and of *Listeria* phage with higher/older metabolic age. However, none of the associations to metabolic age were replicated in the validation set. No antibody responses were significantly associated to methylation age or sj-TRECs.

To account for the co-occurrence of antibodies against CMV with multiple other antibody responses (13), we conducted additional analyses by controlling for CMV serostatus. To determine the CMV serostatus of individuals, we utilized PhIP-Seq data and leveraged serological results from a subset of participants in a different cohort for which PhIP-Seq information was available (25) (see **Methods**). As prediction of CMV serostatus using this model showed high accuracy (median fold accuracy of 0.966), we applied it to predict CMV serostatus in LLD samples.

After accounting for predicted CMV serostatus, we re-evaluated our training and validation associations after accounting for predicted CMV serostatus [Figure 2A] (**Sup. Table 2.2**). This revealed two significant associations to shorter telomeres, present both train and test sets, of two overlapping peptides belonging to the *Bacteroides* fimbrillin family protein (known to constitute bacterial pili) and NK-cell/fully differentiated T-cells (CD45RA+CD57+) TL [Figure 2B]. This protein is important for bacterial adhesion to mucosa, and is well known for its role in pathogenesis of bacteria such as *P. gingivalis* (26), although our results point to fimibrilin peptides from commensal microbiota. The two peptides are highly correlated with each other (phi=0.82), and are more often observed in (predicted) CMV positive individuals (phi=0.26-0.27). In contrast, antibodies against rhinoviral peptides exhibited a significant association with longer telomere length in naïve T-cells and lymphocytes (as observed previous to CMV serostatus correction) [Figure 2B]. Additionally, in spite of the adjustments made for CMV serostatus, two associations with CMV peptides remained significant (peptides with IDs *twist_48890* and *twist_53233*) [Figure 2B]. These peptides belonged to two distinct epitopes from CMV’s ‘phosphoprotein 150 & Large structural phosphoprotein’. There is a total of 35 peptides belonging to those proteins in the library used. The antibody-bound peptide *twist_48890* was highly correlated with CMV serostatus prediction (phi=0.93) and showed a stronger association with TL than CMV serostatus prediction (95% CI effect: *twist_48890* [−0.86, −0.62], CMV prediction [−0.83, −0.58]). On the other hand, *twist_53233* was less strongly correlated with CMV infection prediction (phi=0.58) and showed independent effects on TL (multivariable model, P_CMV_prediction_=2.63×10^−11^, P_twist_53233_= 2.21×10^−5^). This suggests that not all individuals with CMV infection develop an immune reaction against these specific peptides, and those who do tend to have shorter telomeres.

### CMV serostatus relates to NK-cell/fully differentiated T-cell TL and shows sex-specific effects

To follow-up the association between antibody-bound peptides from CMV and biological aging, we used the predicted CMV serostatus (rather than the individual antibody-bound peptides) to perform univariate association with TL and other aging markers [**Sup. Table 3A**]. By utilizing the CMV serostatus prediction, we confirmed our previous observation of an association of antibody-bound peptides of CMV with TL [**Fig 3A**] and a lack of significant associations with other biological aging biomarkers. NK-cells/fully differentiated T-cells (CD45RA+CD57+) had the strongest association with CMV (ordinary least squares (ols), effect=−0.52, P=3.09×10^−22^), but we also observed significant associations in naïve (ols, effect=−0.17, P=1.1×10^−3^) and memory T-cells (ols, effect=−0.22, P=6.4×10^−5^) and lymphocytes (ols, effect=−0.29, P=2.37×10^−8^). T-cells and NK-cells are known to play an important role in controlling CMV infection (27). Some specific NK-cell populations, such as CD57+NKG2Chi-expressing populations or terminally differentiated CD56^dim^CD16− cells (28), are known to expand in people with CMV infection. These expanded populations show memory-like features (29), which could potentially lead to a decrease in the average TL of the NK-cell population, providing a potential explanation for the association we observe. On the other hand, senescent T-cells also tend to accumulate with CMV infections (30).

**Figure 3.**
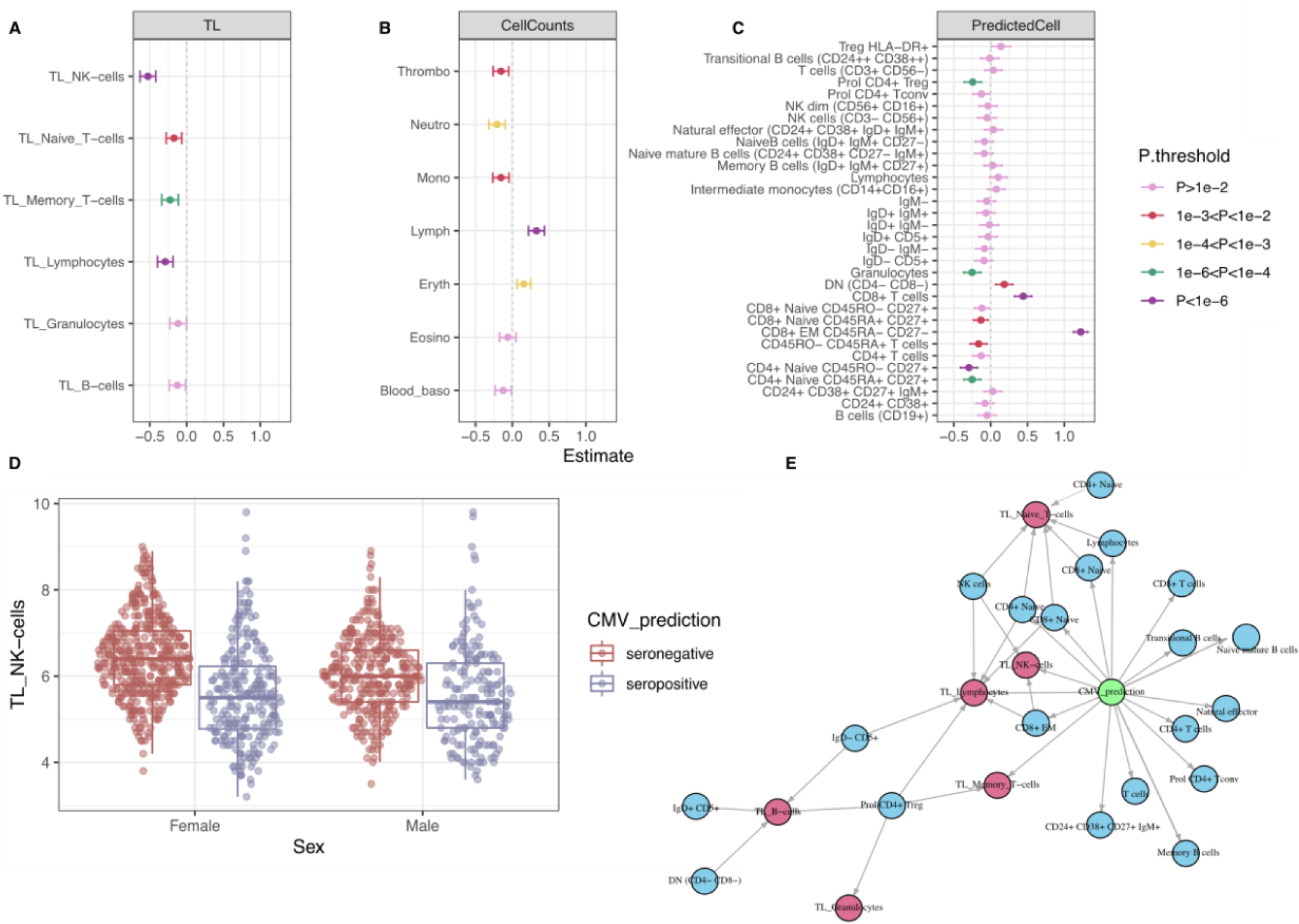
Associations of CMV prediction with cell counts and telomere length. **A–C**, Linear association of CMV prediction (binary) with (**A**) telomere length (TL), (**B**) measured blood cell types, and (**C**) RNA-seq deconvoluted predicted cell counts. Estimated effect values, accounting for age and sex, are displayed. Error bars represent the 95% confidence interval of the estimated effect. **D**. TL differences between men and women in the NK-cell/fully differentiated T-cell population. CMV interacts significantly with sex. **E**. Mediation network. CMV infection effect (green circle) on TL (red circles) is partially mediated by changes in predicted cell counts (blue circles). Effects are estimated using lasso regression. Edges represent absolute effect sizes above 0.01.

In addition, we observed that CMV status acts as a confounding factor for correlations between different biological aging biomarkers. Particularly, in the correlation between the TL of NK-cells/fully differentiated T-cells and all other cell types becomes decoupled in CMV seropositive individuals [**Sup. Table 3B**], as the TLs of NK-cells/fully differentiated T-cells (CD45RA+CD57+) of CMV seropositive individuals are the most reduced among all the TLs.

Furthermore, in a subsequent CMV-sex interaction analysis, we observed a significant decrease of TL in NK-cells/fully differentiated T-cells of women infected with CMV as compared to men (ols, effect=−0.28, P=7.6×10^−3^) [**Sup. Table 3C**]. This indicates that, on average, CMV infection tends to have a greater impact on TL in women compared to men [**Fig 3D**]. Previous studies in this cohort have shown differences in NK-cell/fully differentiated T-cell TL between men and women (mean difference in women=0.2, P=3.45×10^−4^) (16). However, when stratified by CMV infection, the sex differences in TL of CMV-infected individuals are no longer significant (mean difference in infected individuals=0.018, P=0.864; mean difference in non-infected individuals=0.346, P=3.14×10^−8^). This observation could not be attributed to the differences in CMV seropositivity prevalence between the sexes as CMV seroprevalence was similar in males and females (Chi-squared test, P=0.61). In conclusion, while telomeres in women are significantly longer than in men overall, this difference is not significant in CMV-infected individuals due to a larger decrease of TLs in CMV-positive women compared to CMV-positive men.

It was previously reported that women exhibit stronger reactivity against CMV, characterized by increased cytokine secretion (31). This could potentially be linked to a more pronounced reduction in TL among women. To explore this hypothesis, we tested for sex-specific CMV effects in 14 circulating cytokine concentrations, however we did not identify significant associations (FDR_1000permutations_>0.05) [**Sup. Table 3D**].

To further investigate sex-specific differences related to CMV infection, we examined the antibody titers for CMV peptides by comparing the normalized read counts of immuno-precipitated CMV peptides (as a proxy for antibody titers, see **Methods**) from CMV seropositive men and women. Here we observed that women had, on average, a higher number of reads for all CMV peptides (76/77 peptides with higher numbers in women, 45/77 under FDR<0.05) [**Sup. Table 3E**], suggesting a stronger adaptive immune response to CMV. In addition, we observed a similar, albeit not significant, trend in an independent cohort with CMV serology (300BCG, described in **Methods**) (effect_women_ on CMV titers (IU/ml)=1.08, P=0.278).

In the same independent 300BCG cohort, we used qPCR measurements of average blood cell TLs to replicate our association with CMV infection. Despite observing an average decrease of TL in individuals with CMV-positive ELISA, this difference did not show strong statistical support (effect CMV seropositive on TL=−0.69, P=0.3). Sex-specific effects of CMV on TL were also not observed in the 300BCG dataset (interaction effect in women=−0.05, P=0.96). These results might be related to the cell-specific nature of CMV–TL changes: when averaged across different blood cell populations, as is often done through qPCR measurements, the effect might be masked.

In addition to the associations with CMV, we had identified a positive association between anti-rhinoviral responses and TL. While there was a higher anti-rhinoviral prevalence in younger participants, the positive relationship between rhinovirus and TLs could be replicated in age-matched rhinovirus-positive and -negative participants (multivariable model with age, CMV and *twist_35344* on overall TL, effect_twist_35344_=0.21, P_twist_35344_=2.5×10^−4^). In addition, we observed that having a greater breadth of positive anti-rhinoviral peptides was more strongly associated with longer TL than individual peptide reactivity [**Sup. Table 4**] (see **Supplementary** for more information).

### Blood cell-type composition is associates with CMV and partially explains the TL changes

Next, we investigated the association between predicted CMV status and various blood cell types from 1,408 participants [**Sup. Table 3A**]. Using clinical measurements of major cell types in these individuals, we found a strong positive association between CMV infection and lymphocytes (ols, effect=0.32, P=1.77×10^−9^), as well as negative associations with neutrophils (effect=−0.208, P=1.6×10^−4^) and erythrocytes (effect=0.156, P=7.42×10^−4^) [**Fig 3B**]. To further explore the specific lymphocyte groups affected by CMV infection, we used deconvoluted cell proportions derived from bulk-blood RNA-seq data in 1,173 participants [**Fig 3C**]. This deconvoluted dataset includes estimated cell proportions for 32 cell types, to which we applied a log-ratio transformation to account for cell proportion interdependencies introduced by data compositionality. Here we observed strong positive associations between CMV infection and CD8+ TEM CD45RA-CD27-(effect=1.22, P=5.66×10^−110^) and CD8+ T-cells (effect=0.44, P=7.38×10^−14^), and negative associations with the naïve subtypes CD45RO-CD27+ (effect=−0.295, P=2.45×10^−7^) and proliferative CD4+ T-reg (effect=−0.243, P=3.73×10^−5^), as well as with granulocytes (effect=−0.252, P=1.9×10^−5^). Positive associations with memory cells and negative associations with naïve cell types are generally well-established patterns in CMV infection (32–34).

Since blood cell-type composition might be related to average TLs if the cell populations have heterogenous TLs, we examined whether the observed association between CMV and TL could be explained by changes in the composition of blood cell type. To investigate this, we conducted a mediation analysis and found that the measured cell populations partially mediated the changes in TL in certain cell types. However, a substantial proportion of the variability remained unexplained, which suggests changes in other unmeasured cell types or mechanisms through which CMV can affect TL independently of changes in (predicted) cell-type composition [**Fig 3E**]. For example, in the case of NK-cells/fully differentiated T-cells, the major mediator of TL was the abundance of CD8+ EM cells, which accounted for 36% (95% CI, 15.4%–53%) of the CMV effect on the TL if NK-cell/fully differentiated T-cells. This indicates that a significant portion of the CMV effect on TL was not dependent on changes in predicted cell counts.

Interestingly, we observed opposite associations between cell-type composition and anti-rhinoviral responses than those observed for CMV. Similar to CMV, such cell changes only partially mediated the rhinoviral effects on TLs (see **Supplementary** for details) [**Sup. Fig 1**].

### CMV is associated with gene expression changes in specific cell types

Previously we found an association between CMV seropositivity and predicted cell-type composition. However, such analysis may be prone to biases related to cell prediction algorithms using bulk RNA-seq. It also does not identify expansions of specific cellular subpopulations and overlooks transcriptional effects at the cellular level that are unrelated to cell expansions. To shed light on the molecular processes associated to CMV seropositivity in specific blood cell populations, we used 3’-end single-cell RNA-sequencing (scRNA-seq) data previously generated on cryopreserved, unstimulated PBMCs from a subset of the LLD participants, Oelen2022 (n=94, CMV seronegative=59, CMV seropositive=35) (35). While the scRNA-seq data collection was posterior to PhIP-Seq (5 years on average), previous literature indicates that infection with CMV late in life is rare (34). This is consistent with our observations [Figure 1A], where only 2 out of 325 individuals with data from two timepoints (3.5-years apart on average) were predicted to be CMV-negative at baseline and CMV seropositive at follow-up. We therefore concluded that we can use CMV status inferred from PhIP-Seq data to label scRNA-seq data from the same donor despite the difference in collection time. In addition, we accounted for potential technical differences between V2 and V3 10X Chromium Single Cell 3’ chemistries in all of our analyses (see **Methods)**.

First, we associated CMV serostatus with blood cell composition predicted from scRNA-seq. These results reproduced our previous associations between CMV serostatus and blood cell counts (either measured with EDTA Sysmex **[Fig 3A]** or deconvoluted from bulk RNA-seq **[Fig 3B]**), including the positive association between CMV and both CD8+ T-cells and its subpopulation CD8+ TEM and the negative association with CD4+ naïve T-cells and a depletion of regulatory T-cells (T-reg). In addition, the single-cell data allowed us to observed an additional expansion of CD4+ cytotoxic T lymphocytes (CTL) in CMV seropositive individuals [**Fig 4A**] **[Sup. Table 5A]** (see **Supplementary Materials** for more information).

**Figure 4.**
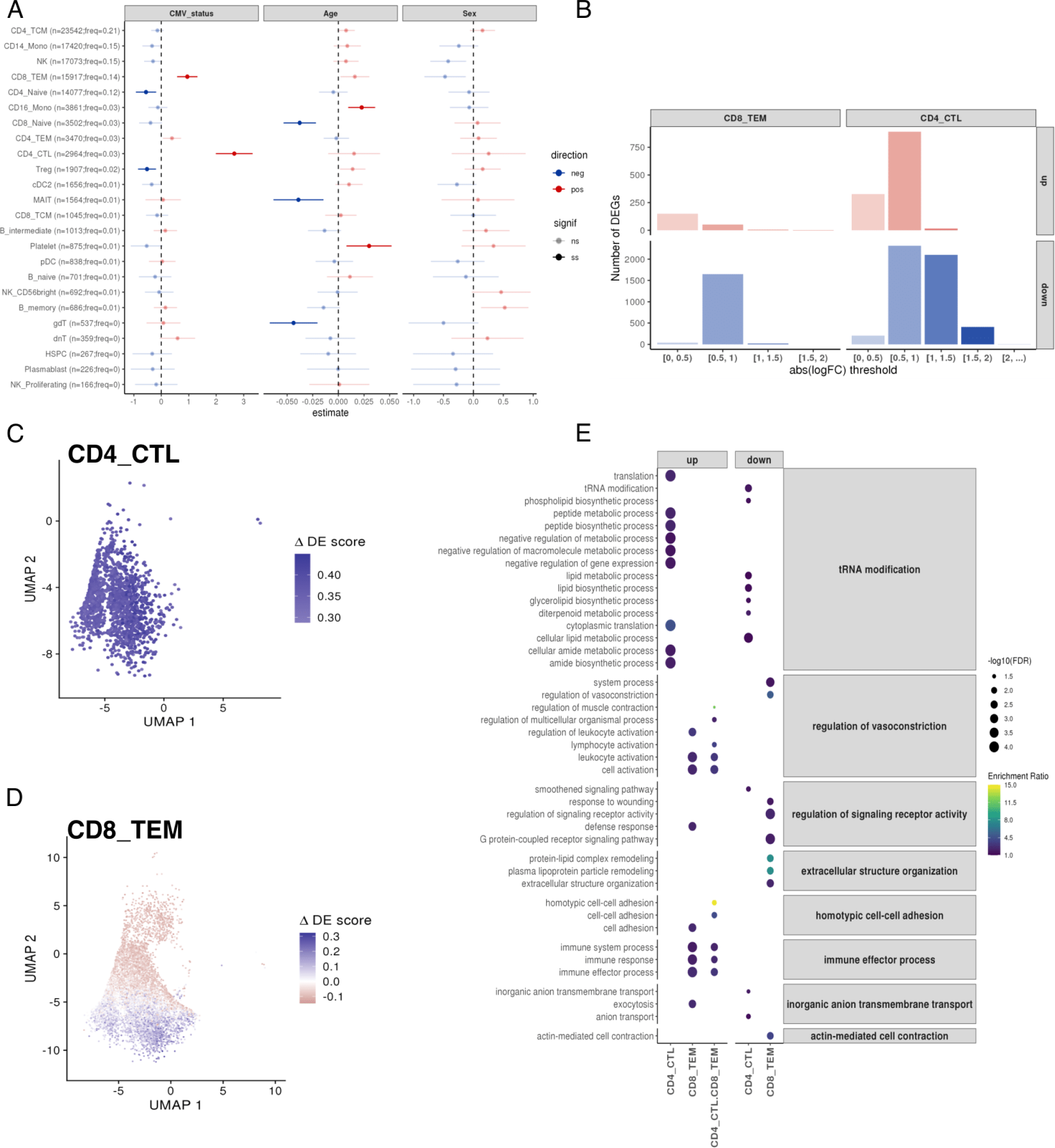
CMV seropositivity associates with both cell composition and cell-type-specific gene expression changes. **A.** Forest plot showing the linear association of CMV serostatus, age and sex with high-level (Azimuth’s l2) cell-type-proportions from Oelen2022 (35) scRNA-seq data. Estimated effect values, accounting for both biological (sex and age) and technical (10X Chromium Single Cell 3’ chemistry and experimental batch) covariates, are displayed. Error bars represent the 95% confidence interval of the estimated effect. The absolute number and relative frequency of each cell type is shown. **B.** Bar plot of the number of CMV-differentially expressed genes (DEGs) across cell types split by the direction (up- or down-regulated) and magnitude of the effect size (x-axis). **C–D.** UMAPs showing the ΔDE (differential expression) scores using data from CD4+ CTL (**C**) and CD8+ TEM (**D**). **E.** Enriched gene ontology (GO) terms, using the biological process pathway database, from three different CMV-DEG sets using the Oelen2022 data: CD4+ CTL-DEGs, CD8+ TEM-DEGs, and DEGs shared between CD4+ CTL and CD8+ TEM. GO terms are grouped based on their semantic similarity to simplify the redundancy of GO sets. Dot color indicates enrichment ratio. Dot size indicates statistical significance.

CMV may indirectly affect TL by affecting sub-cell-type composition. Previously, we observed an association between CMV and TLs from mature NK/T-cells (expressing CD45RA+CD57+). We wondered if different sub-cell-types within CD45RA+CD57+ may be amplified with CMV infection, thus causing changes in the population’s TL. Investigation of this cell population is not straight-forward, however. While we could use the expression of the CD57-encoding gene (*B3GAT1*) to determine CD57-positivity, the 3’-biased mRNA capturing of 10X’ scRNA-seq data did not allow us to distinguish the expression of CD45RA/RO **[Sup. Fig 2B**]. We observed enough cells with expressing *B3GAT1* for analysis in NK-cells, CD8+ TEM and CD4+CTL **[Sup. Fig 2C**] and saw an increase in the abundance of *B3GAT1*+ cells (centered-log-ratio (CLR) transformed) in CMV-positive individuals (effect=0.9, P=3.75×10^−4^). However, the association between CMV and different cell populations within *B3GAT1*+ cells was not homogeneous (likelihood ratio test model with CMV-cell-type interaction vs base model, P=6.88×10^−4^). To understand whether B3GAT1+ cell amplification differs between cell types, we conducted stratified analyses testing for the effect of CMV in each cell type expressing *B3GAT1*. We found that CMV seropositivity significantly increased CD8+TEM B3GAT1+ (effect = 1.53, FDR=4.95×10^−4^) and CD4+ CTL B3GAT1+ (effect=1.37, FDR=2.89×10^−4^). On the other hand, CMV infection showed a non-significant negative trend in NK B3GAT1+ (effect =−0.3, FDR=0.48) **[Sup. Table 5A]**. These results suggest that, among the cell populations in which TL was greatly influenced by CMV seropositivity, NK-cells were not largely affected whereas both CD8+ TEM and CD4+ CTL were increased. This differential amplification of cell populations might be related to the observed TL shortening. However, it is important to highlight that only cells among those that also express CD45RA could be measured by our Flow-FISH probe, and thus the results might misrepresent which populations are expanded in the final measurement. In addition, we checked for expansion of NK-cells expressing NKG2C (i.e. with transcription of the gene *KLRC2*), a cell population usually associated with CMV infection (36), however we did not identify a significant increase of that subpopulation in CMV seropositive individuals (effect=0.54, P=0.17).

Next, as we wondered if CMV infection by itself may alter the transcriptional landscape of immune cells, we performed pseudobulk differential gene expression (DGE) analyses between CMV seropositive and seronegative individuals for each of the predicted cell types, while controlling for age, sex and technical factors (see **Methods**) [**Sup. Table 5B**]. This identified gene expression changes with CMV serostatus in only two T-cell subpopulations: CD8+ TEM and CD4+ CTL cells. We found 7,877 unique differentially expressed genes (DEGs) at FDR<0.05, showing a significant predominance (estimate=0.86, exact binomial test P-value< 2.2×10^−16^) of down-regulated genes (n=6,737) compared to up-regulated genes (n=1,355) [**Fig 4B**]. Notably, most of the DEGs were CD8+ TEM- or CD4+ CTL-specific, with only a small fraction of both up-(88 shared-DEGs out of 1,355 up-DEGs; 0.07 Jaccard Index) and down-DEGs (191 shared-DEGs out of 6,546 down-DEGs; 0.03 Jaccard Index) shared among these subpopulations. Interestingly, while the changes in transcription in CD4+ CTL cells were seen in all cells in the population [**Fig 4C**], the changes in CD8+ TEM cells were attributed to a subpopulation of cells with a higher differential expression score (ΔDE) (this score considers the combined expression pattern of up- and down-regulated DEGs, see **Methods**) [**Fig 4D**]. The positive association we had previously observed between the proportion of CD8+ TEM B3GAT1+ and CMV seropositivity made us wonder whether the population of CD8+ TEM cells enriched in ΔDE could contain such a subpopulation [**Sup. Fig 3A–D**]. Indeed, we found higher ΔDE scores among CD8+ TEM B3GAT1+ cells compared to the B3GAT1-cells (Wilcoxon rank sum test P-value<2.2×10^−16^) [**Sup. Fig 3E–F**]. Thus, we expanded our DGE analysis to the subset of CD8+ TEM and CD4+ CTL that were B3GAT1+ and B3GAT1-in order to define subpopulation-specific transcriptional changes with CMV seropositivity. Although limited by the reduced sample size of the B3GAT1+ subpopulations (375 cells in CD8+ TEM B3GAT1+ vs. 15,917 cells in CD8+ TEM, 85 cells in CD4+ CTL B3GAT1+ vs. 2,964 cells CD4+ CTL), we identified 73 DEGs in CD8+ TEM B3GAT1+ cells [**Sup. Table 5C**]. CD8+ TEM B3GAT1+ DEGs largely overlap with the ones identified in CD4+ CTL cells (40 out of 42 up-regulated CD8+ TEM B3GAT1+ DEGs and 17 out of 31 down-regulated CD8+ TEM B3GAT1+ DEGs). This indicates a cytotoxic-like response of this CD8+ TEM B3GAT1+ subpopulation that is expanded by CMV seropositivity [**Sup. Fig 3G-H**].

To follow-up on this, we assessed whether our reported DEGs belonged to similar functional pathways, thereby highlighting the biological interplay between CMV seropositivity and gene expression [**Fig 4E**]. Here we observed the enrichment of several pathways both for up- and down-DEGs for CD4+ CTL and CD8+ TEM [**Sup. Table 5D**]. This included the enrichment of negative regulation of metabolism and gene expression in CD4+ CTL and an enrichment of leukocyte activation and immune effector process in CD8+ TEM cells, among others [see **Supplementary** for a more extensive discussion].

### Viral infections associate with circulating inflammation-related cytokine concentrations

We then wondered if, in addition to affecting biological aging, previous and chronic exposures could be related to systemic inflammation. The mechanisms driving such changes might be related to trained immunity, a set of epigenetic changes in innate immune cells triggered by past immune encounters. To explore this, we utilized the complete dataset of 2,815 peptides (bound in 5–95% of individuals) with antibody information together with measurements of 14 blood cytokine and adipokine concentrations obtained through ELISA from 989 participants.

For the multivariate analysis, we employed sparse canonical correlation analysis (sparse-CCA), a powerful technique that has been successfully utilized to integrate diverse biological layers (37,38). This approach allowed us to identify coordinated patterns of associations between antibody profiles and inflammation-related markers. We also performed univariate analysis using HAllA, as in our previous analysis. This strategy enabled us to explore individual associations between specific antibodies and inflammation markers, while considering both training and validation datasets for robustness and generalizability [**Fig 1B**].

Our analysis revealed a positive association between the anti-inflammatory cytokine Interleukin-18 Binding Protein (IL-18BP) and viral signals that was particularly driven by adenovirus C (Pre-histone-like nucleoprotein, rho_partial_Validation_=0.239) and EBV (two associations with nuclear antigen 1 (rho_partial_Validation_=0.295 and 0.269) and one with nuclear antigen 2 (rho_partial_Validation_=0.187)). These associations were consistently observed using both the sparse-CCA [**Sup. Table 6.A-B**] and HAllA [**Sup. Table 6.C-D**] methodologies, underscoring their robustness and reliability [**Fig 5A, 5C**]. Furthermore, prediction models utilizing PhIP-Seq data demonstrated that individual circulating IL-18BP concentrations could be better predicted using antibody responses than by a simple model based on age and sex alone, further supporting the relationship between IL-18BP and antibody profiles [**Fig 5D**].

**Figure 5.**
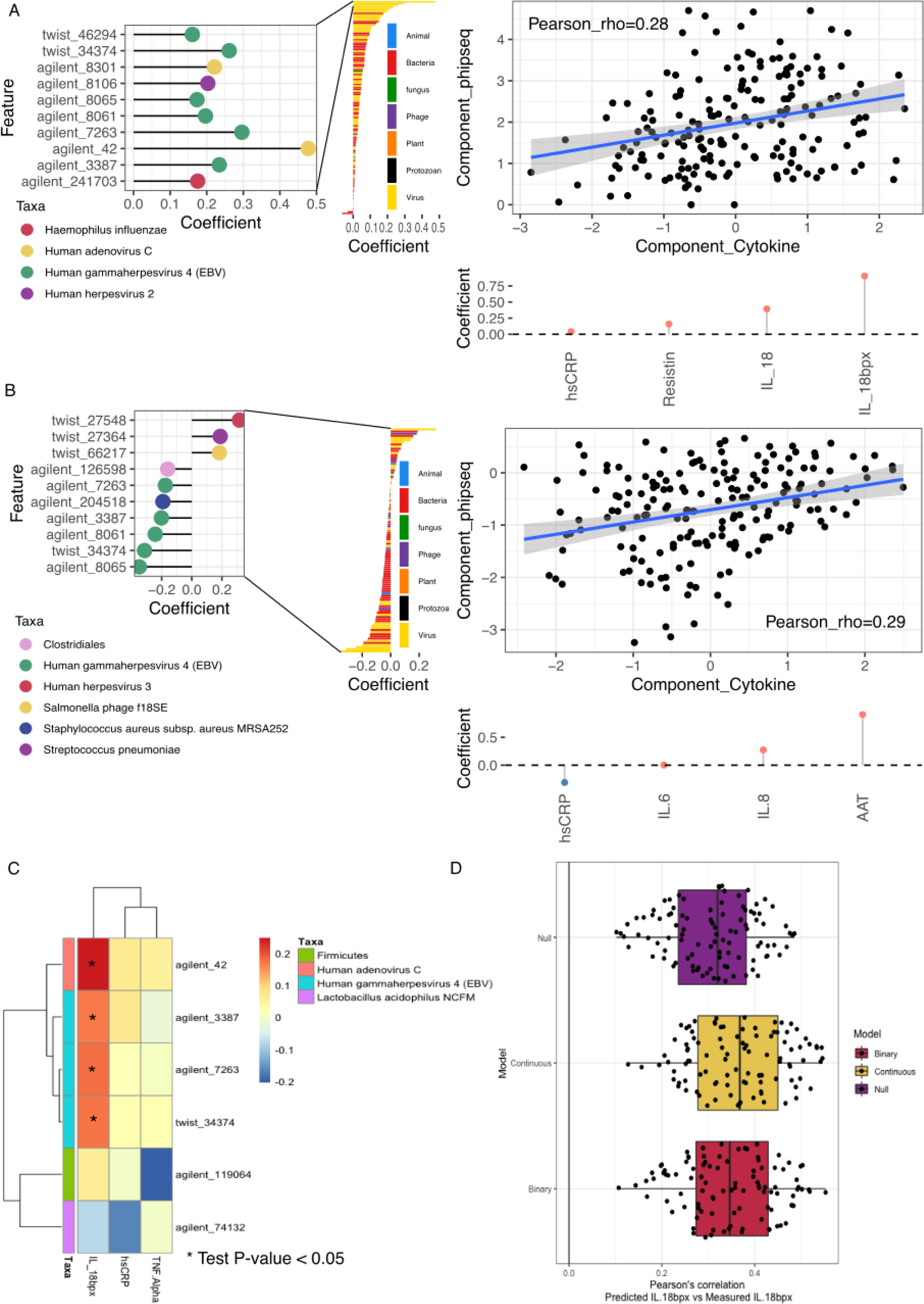
Circulating cytokines and antibody epitope reactivity. **A-B.** Sparse-CCA components 1 (**A**) and 4 (**B**), which associate a component loaded by presence/absence PhIP-Seq profiles (Y-axis) and circulating cytokine concentrations (X-axis). Right panel shows the correlation between PhIP-Seq and cytokine component in test-data. Left panel shows top antibody-bound peptide loads in the Phip-Seq component (Y-axis in right panel). Bottom panel shows the top cytokine component loads in the Cytokine-component (X-axis in right panel). **C.** HAllA heatmap of associations. X-axis shows the cytokines that had at least one significant association in the training data. Color indicates the Spearman’s correlation in the training data. * indicates a partial Spearman’s correlation P-value < 0.05 in the validation data. Y-axis displays the different peptides with annotation indicating the organism where the peptide originates. **D.** Prediction results for IL-18BP using an L2-regularized linear model with all peptides, using presence-absence scores (binary), normalized read counts (continuous), or prediction using ordinary least squares of age and sex. Each dot represents the Pearson’s correlation of the held-out validation data’s prediction and measurement. Estimations include rho values for 10-fold cross-validation repeated 10 times.

The balance between IL-18BP and IL-18 is important in proinflammatory responses (39). IL-18 stimulates proinflammatory cytokine production in NK cells and CD4+ T-cells (40), and IL-18BP concentrations have been observed to be negatively correlated with other circulating proinflammatory cytokines in the population (41). While high concentrations of IL-18 have been observed during acute EBV infection (42), we identified that individuals with chronic EBV infection display higher concentrations of IL-18BP. Similarly, individuals with antibodies against adenovirus C also exhibited higher concentrations of IL-18BP. It is worth noting that some viruses are known to express IL-18BP-like molecules as a means to dampen the host immune response (43), and it is possible that similar strategies inducing IL-18BP expression exist during viral infections to promote tolerogenic reactions.

In addition to the associations involving IL-18BP, our analysis identified other signals. In sparse-CCA component 4 (rho=0.287, P=7.36×10^−5^) [**Sup. Table 6.A-B**] [**Fig 5B**], we observed a negative association between EBV and concentrations of α-1-Antitrypsin (AAT), an endogenous protease inhibitor that alleviates tissue damage produced during inflammation, and other proinflammatory cytokines such as IL-6 and IL-8. Conversely, AAT concentrations were positively associated with human herpesvirus 3. On the other hand, high-sensitivity C-reactive protein (hsCRP), a biomarker for inflammation, was increased in individuals showing more EBV antibody-binding. Sparse-CCA component 4 was dominated by EBV peptides but was also influenced by other bacteria and phages, including *Staphylococcus aureus subsp. Aureus MRSA252* and *Streptococcus pneumoniae*, as indicated by their loadings (0.192 and 0.189, respectively).

Altogether, these results suggest that chronic infections, such as EBV, may have an effect on circulating cytokine concentrations of the general population.

## Discussion

In this study we performed a comprehensive analysis linking past and current exposures with several markers of biological aging and systemic inflammation. By determining the presence of antibody responses against common pathogens and gut microbiota commensals, we were able to define previous and chronic exposures that were recognized by the immune system and generated an adaptive immune response.

We identified that TLs of immune cells display important relationships with exposures. Most of this signal was driven by a relationship between CMV infection and shorter TLs of NK-cells/fully differentiated T-cells (CD45RA+CD57+). While a relationship between CMV infection and TL reduction has been described previously (44), our cell-population-specific approach enabled by Flow-FISH allowed us to fine-map this response to specific cell populations. CD45RA+CD57+ cells might play a role in CMV homeostasis, where CD57+ NK-cells are observed to be an expanded NK-cell memory subtype triggered by CMV infection (29,45). On the other hand, expression of CD45RA+CD57+ is also seen in senescent T-cells (46). CMV is known to drive the expansion and accumulation of memory (known as memory inflation) and senescent cells (34,47). Such cells generally have shorter TLs since they have undergone cell expansion and telomerase activity is progressively lost in differentiated cells (44). We postulated that CMV-induced changes in sub-cell-type composition might drive part of the observed CMV associations with TL, and our analysis of cell counts using deconvoluted data from bulk-blood RNA-seq supported that CMV was associated with an expansion of memory cells, in particular CD8+ TEM. However, this expansion could only partially explain the observed differences in TL, suggesting that other cell-composition-independent mechanisms might be at work. To validate this at higher resolution, we used scRNA-seq data and observed that the populations of CD8+ TEM cells and CD4+ CTL cells expressing CD57+ expanded with CMV infection, while NK-cells expressing CD57+ did not. Previous work has pointed to expansion of CD8+ EMRA T-cells (48) (expressing CD45RA+ and commonly CD57+) after CMV infection, which also supports this observation. The differential expansion of senescent T-cells expressing CD57+ and CD45RA+ might drive the telomere changes if their average TLs were shorter than those of NK-cells expressing CD57+ CD45RA+. Thus, to fully comprehend if this association is solely driven by that expansion, TLs of CD57+ CD45RA+ NK-cells and T-cells ought to be measured separately.

On the other hand, our absent of association between expansion of NK-cells and CMV infection contrast previous literature where the specific expansion of the subpopulation NKG2C is commonly observed (49,50). Such population of NKG2C, is known to encode CD57+ (49,51) and should be part of the population analyzed through our single-cell analysis. Interestingly, NKG2C cells have been shown to play a fundamental role in protection EBV-infected individuals to develop MS (52). CMV infection is believed to have a protective effect on multiple sclerosis, and this mechanism could be mediated by amplification of NKG2C populations that kill T and B-cells targeting the EBV-EBNA-1 antigen and that cross-react with the glial cell adhesion molecule (52).

We also identified positive associations between rhinoviral immune responses and TLs. Telomerase is known to be activated during acute infections (53). The fact that no flu-like pathogen other than rhinoviruses is linked with TL might be related to the higher infection rates for rhinoviruses during sampling seasons. Data from the National Institute for Public Health and the Environment (RIVM) from the Netherlands indicates that the prevalence of rhinovirus and rotavirus were the highest among respiratory viruses during the sampling for the study (weeks 18–44 from 2012) (data not shown). Another possible explanation is that individuals with longer telomeres might have higher potential to produce antibody responses against rhinoviruses. In a previous study, both longer telomeres in PBMCs and antibody titers against rhinovirus were observed to reduce the odds of rhinoviral infection, which might relate to both higher rhinoviral antibodies and TL (54).

Finally, we uncovered a relationship between EBV and adenovirus C and higher circulating IL-18BP concentrations. Given the anti-inflammatory effect of IL-18BP, these results seem to suggest a tolerogenic response to these infections, which is also supported by the observed negative associations to IL-6, IL-8 and AAT. IL-18BP has been shown to have a role in other viral diseases, for example higher concentrations of IL-18BP are present with higher severity of both Sars-coV-2 (55) and dengue (56) infections. Those higher concentrations of IL-18BP are normally accompanied by increases in IL-18, but generally similar concentrations of free (not bound to IL-18BP) IL-18. Elevated IL-18BP and IL-18 concentrations are also a hallmark of inflammatory conditions such as inflammatory bowel disease (57). Our results also highlight a sparse-CCA component with both IL-18BP and IL-18 that is highly associated with viral peptides from EBV and adenovirus C. Overall, an increase of IL-18BP might be a hallmark of systemic inflammation as a compensatory mechanism by the immune system to avoid overactivity.

All in all, in this study we have explored the links between previous or chronic exposures that induce adaptive immunity and widespread markers of biological aging, blood cell-type composition and inflammation. Our results highlight the impact of chronic infections in biological aging and systemic inflammation. However, we also found signals related to frequent challenges, including adenovirus and rhinovirus, that might similarly leave an imprint on the immune system (adenoviruses) or, conversely, be a challenge to which the immune system should be able to respond to well (rhinoviruses). These results provide a comprehensive overview of how immunological challenges may affect healthy aging by modulating systemic inflammation and TL. In particular, we point to CMV as the immune exposure most associated with telomeric changes in blood cells, thereby further contributing to evidence for CMV’s important role in cellular senescence among all other human viruses (47), although other intriguing associations also emerged with responses against rhinoviruses and bacterial fimbrillin that require further validation. At the same time, we identify circulating cytokine signatures related to EBV and adenovirus C immune responses that might point to a mechanism of action of the virus or the host to reduce systemic inflammation. All in all, our study identifies and describes the effect of common commensal and pathogenic bacteria and viral exposures on the immune system of the general population.

## Methods

### Cohort information

We used data from the Lifelines cohort. Lifelines is a population-based cohort study that collects health and health-related behaviors from individuals from the North of the Netherlands (58). For a subcohort of Lifelines, Lifelines-DEEP (LLD), additional molecular information is available. We used data for 1,437 LLD participants (57% women, age=44.6±14 years, 340 with follow-up 4-years apart). For all participants we have self-questionnaires regarding lifestyle and health and anthropometric and blood parameters. For different subsets of this cohort, different molecular data is available.

### Data generation

#### PhIP-Seq libraries and enrichment

PhIP-Seq profiles were previously generated (13) as described earlier (11,12), following a previously described protocol (59). Two peptide libraries were profiled. These libraries used different chemistry and contained peptides of different origins: the 244,000 peptide library manufactured by Agilent Technologies, Inc. (here named ‘Agilent’ for brevity) is enriched in bacterial peptides (11) and the oligopeptide library of 100,000 variants manufactured by Twist Bioscience (‘Twist’ library) is enriched in known immune epitopes (12). Antibody-bound peptide presence/absence enrichment was previously determined (13).

PhIP-Seq data is not directly interpretable using sequencing read counts. On the one hand, raw reads are not comparable between samples as the sequencing experiment yields compositional data. On the other hand, different phages might precipitate without an actual antibody attachment. To account for these possibilities, PhIP-Seq experiments include a sequencing run without immunoprecipitation, which yields the “input levels” for a certain peptide, i.e. the number of reads from a peptide that are obtained without immune-precipitation. Antibody reactivity can then be induced by comparing read levels of peptides with the same input levels within the same individual, for which data compositionality is not an issue. Read numbers from a certain input level are assumed to follow a generalized Poisson distribution, and P-values can be obtained as previously described (11). Bonferroni-corrected P-values below a threshold of 0.05 were used to define antibody-bound peptides in a binary fashion.

In addition, to obtain continuous scores that can act as a proxy for antibody titers, we normalized transcript per million abundances, obtained using Salmon (60), using log-ratios. Log-ratios are commonly used in compositional data analysis (61). The abundances of a specific peptide (plus a pseudocount) are then expressed relative to a denominator that is thought to be constant in different samples. In this case, we used a median-of-ratios strategy to normalize the data. First, we computed the geometric mean among samples in two overlapping *Staphylococcus aureus* peptides representing protein A (TPA B-domain-containing protein) (*agilent_221096* and *agilent_133222*, respectively). Such protein binds to the fragment crystallizable region of the antibodies, leading to widespread immunoprecipitation events not caused by direct antibody recognition, as previously described (11). Reactivity of these two peptides were predicted in all but three samples, which were removed from the continuous analysis as the denominator is not expected to be constant in such samples. Second, per sample, we divided the values of *agilent_221096* and *agilent_133222* by their inter-sample peptide-specific geometric mean. Finally, we computed the mean value of the normalized *agilent_221096* and *agilent_133222* abundances per sample, yielding a sample-specific normalization factor. We used this factor as the denominator in the log-ratio transformation for all other peptides.

This data yields a continuous score that is independent from thresholds, but it comes with the caveat that a certain amount of baseline reads are expected even without immunoprecipitation and this might introduce a certain level of noise into the associations. Due to this issue, read abundances are not directly comparable between peptides.

#### Proteomics and cytokine measurements

Proteomics profiles were generated using the Olink^®^ CVD panel III for the 1,433 LLD participants with PhIP-Seq profiles (14). This panel includes 92 proteins previously linked to cardiovascular disease. The protein CCL22 was removed from the analysis based on indications from Olink. Adipokines and cytokines were measured previously in the LLD cohort (62). Cytokines included IL [interleukin]-1β, IL-6, IL-8, IL-10, IL-12p70 and TNF-α (tumor necrosis factor-α). Leptin, adiponectin, IL-18, IL-18BP, resistin and AAT (alpha-1 antitrypsin) were measured using ELISA kits (R&D Systems, Minneapolis, MN).

#### Bulk RNA-seq of participant’s blood

Whole-blood RNA-seq data were generated previously(63). Paired-end reads were mapped to the human genome using STAR (64), allowing for a maximum of seven mismatches. HTSeq-count (65) was used on uniquely mapping reads to quantify transcripts. We used blood cell proportions previously predicted from whole-blood bulk-RNA profiles using Decon2 (66). The main advantage of Decon2 is that the prediction does not depend on measuring gene expression in purified cells to determine gene signatures. Using Decon2, we predicted the proportions of 34 cell populations. In addition to the predicted cell counts, we measured lymphocytes, erythrocytes, monocytes, neutrophils, thrombocytes and eosinophils in participant’s blood with EDTA Sysmex. Additive log-ratio transformation (ALR) was performed on predicted cell proportions, using the proportions of monocytes as denominator.

#### TLs of immune cells

TL measurements for 1,046 LLD individuals were generated previously using Flow-FISH for six blood cell types: granulocytes, lymphocytes, B-cells (CD45RA+CD20+), naïve T-cells (CD45RA+ CD20-), memory T-cells (CD45RA-) and NK-cells/fully differentiated T-cells (CD45RA+CD57+) (16). For some individuals, the TLs of certain cell types were missing (109 missing samples for granulocytes, 9 for B-cells and 17 for NK-cells, with no samples missing TLs for more than one cell type). Missing points were imputed by building a linear prediction model of the TL based on all other TLs. To assess imputation quality, samples with no missing telomere information were split into training data (80%) and held-out test data (20%). We used a linear model (ols) using a specific TL as dependent variable and including all other TLs as features for prediction. Using the trained-fit model, we computed the R^2^ of the prediction in the test. The imputation showed high resemblance to the actual held-out values: R^2^=0.763 for granulocytes, R^2^=0.881 for B-cells and R^2^=0.740 for NK-cells. Thus, we used the model to impute all missing values for the samples not used to build and test the model.

#### Residualization of omics layers

Since neither sparse-CCA or HAllA directly account for possible confounders, we performed a residualization procedure for each omic layer to remove the variability attributed to possible confounders. Covariates that might act as confounders, including age, sex, BMI, blood cell counts, smoking habits and oral contraceptive use, were tested against the distance matrix of each omic layer using permutational analysis of variance (PERMANOVA) implemented in the adonis2 function of R package vegan (v2.6-4). Significant features were then chosen as covariates for residualization. We built independent linear models per feature within an omic layer, including the possible confounders detected by the PERMANOVA analysis as covariates. Residuals of the model were extracted and used as input features for the sparse-CCA and HAllA analyses.

### Dataset split

Before the sparse-CCA and HAllA analyses were performed and the datasets were residualized for possible confounders, participants were randomly split into a training dataset (80% of the data) and a held-out validation dataset (20% of the data). Residualization of datasets was then performed independently in the training and validation sets. Multivariate analyses (HAllA and sparse-CCA) were first performed in the training set and subsequently validated in the validation set.

### Sparse-CCA

To correlate different omic layers, we used a sparse-CCA framework. PhIP-Seq antibody-bound peptides were filtered for a minimal prevalence of 10% in LLD individuals with matching omic layers. In the training set, we performed a grid search (with size and range changed depending on the molecular type analyzed) to optimize the lasso penalty parameters per dataset. A 5-fold cross-validation was performed (R, caret v6.0-90) using the function CCA (R, PMA v1.2.1), searching for a unique component in the output (K=1) and testing all possible combinations of penalties. We assessed the model’s performance using the 1-fold held-out data, for which we projected both omics layers to the trained-found dimensionality and ran a Pearson correlation analysis. Correlation rho were averaged between folds as an assessment of a specific hyperparameter selection. Hyperparameters that achieved a maximum mean rho were selected. Using the complete test-set and the identified hyperparameters, we ran sparse-CCA searching for 10 new dimensions (K=10). Then, to assess the correlation found in the new dimensionality in an unbiased manner, we projected the 20% held-out validation data to the found space and performed Pearson and Spearman correlations between both dimensional projections. The resulting rho and P-value from the validation set were recorded. A P-value<0.05 was considered significant and analyzed in more detail.

### HAllA analysis

Residualized training omics layers were associated with binary antibody profiles using HAllA v 0.8.20. Spearman correlations were used with thresholds set to --fdr_alpha 0.05 and -- fnr_thresh 0.2. We then reproduced the HAllA associations in tests using partial Spearman correlations in non-residualized data while accounting for the covariates of interest.

### PhIP-Seq predictive models

We used continuous and binary (non-residualized) PhIP-Seq data to predict individual cytokine concentrations and TL. For that, we fitted two linear models, an ols using age and sex to predict the dependent variable and a lasso-regularized model including all peptides on top of covariates (R glmnet, v4.0-2). The strength of the regularization parameter λ was chosen from the default grid search from cv.glmnet using 10-fold cross-validation in bases to the obtained root of the mean squared error. To assess an unbiased model performance, a 10-fold cross-validation framework was used. Test fold was used for prediction, and Pearson’s correlation values between predicted and actual values were used to assess model performance. To estimate the variability of such estimates, cross-validation was repeated 10 times, yielding 100 estimates of performance. We then used a t-test to compare estimated predictability between the base and PhIP-Seq models.

### CMV analysis

#### Serostatus prediction

To predict CMV seroprevalence from PhIP-Seq antibody-bound peptide data, we first used a cohort of 497 patients with inflammatory bowel disease with PhIP-Seq profiles (25) where a subset of participants had ELISA seroprevalence information for CMV (n=294 participants, 114 seropositive, 38%). We tested three algorithms in a 5-times 5-fold cross-validation framework: a logistic regression model using the number of positive antibody-bound peptides as the predictor, a logistic regression model with all CMV peptides using a lasso penalty and a logistic regression using as regressor the first Redundancy Analysis (RDA) of PhIP-Seq and CMV seroprevalence component. In all models, age and sex were used as covariates. We used complete PhIP-Seq profiles as a predictor for LASSO, and regularization strength parameter λ was optimized using 5-fold cross-validation. We used RDA to identify the linear combination of the peptide-bound antibody data enrichment that better explains CMV ELISA measurements. We then projected the testing data to the trained RDA space and predicted CMV seropositivity using the resulting RDA-1 component as the regressor of logistic regression. Model accuracies were calculated as the proportion of correct predictions out of all predictions. Statistical testing to compare model performances was done using an ANOVA, followed-up by a post-hoc Tukey test. We selected the top-performing model, which was the logistic regression based on the number of positive antibody-bound peptides, and used it for prediction in the whole LLD dataset. A probability of CMV of at least 0.5 was used to predict CMV seropositivity.

#### CMV associations with cell composition and TLs

CMV predictions were associated with measured blood cell counts, additive log-ratio-transformed predicted blood cell counts (using predicted monocyte proportion as denominator), and TL measurements, all accounting for age and sex. To test for interaction analyses, we included either a CMVxSex or a CMVxAge term in the model. Mediation analysis between CMV (exposure), predicted cell counts (mediators) and TLs (outcome) were run using mvregmed (R regmed v2.0.4), using a grid search of λ parameters between 0.4 and 0.01 (step of 0.01). The best-fitting model was selected based on AIC, and findings were visualized in a network. Individual mediation effects were calculated using mediate v4.5.0

#### CMV-stratified correlation analysis

Here we used the 271 samples for which there were TL measurements, methylation age clocks, metabolic age clocks and qPCR measurements of sj-TRECS. We divided that dataset into participants predicted to be CMV-positive and those predicted to be CMV-negative. We computed the biomarker-to-biomarker Pearson’s correlation coefficient individually in the CMV-positive and CMV-negative samples. To determine whether the observed differences in correlation coefficients were not due to random sampling, we performed Fisher’s Z-transformation by applying the inverse hyperbolic tangent function on the coefficients. Then, we estimated the standard error (SE) for the differences in z-scores using Equation 1.

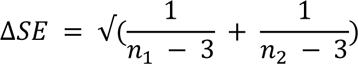

**Equation 1**. SE difference, where *n* indicates the number of samples used to estimate the correlation coefficient.

Next, we divided the difference of z-scores by the estimated SE difference (Equation 1) to obtain a Z-statistic. Finally, we estimated the P-value of the null hypothesis that the correlation coefficients are the same as the probability lower or equal to achieve the estimated Z-statistic on a standard gaussian distribution.

We repeated the above analysis after removal of the “age” effect from the aging biomarkers by regressing out age using a linear model and taking the resulting residuals for correlation.

### CMV and immune single-cell transcriptomics analysis

#### scRNA-seq datasets

We used two previously processed 10X Genomics’ scRNA-seq datasets on cryopreserved PBMCs from a subset of the LLD participants of the present study (n=119): *i)* unstimulated data from Oelen2022 (35) generated using 10X Chromium Single Cell 3’ V2 (n=64, CMV seronegative=43, CMV seropositive=21) and V3 (n=30, CMV seronegative=16, CMV seropositive=14) chemistries and *ii)* unstimulated data from Wijst2018 (67) generated using 10X Chromium Single Cell 3’ V2 (n=25, CMV seronegative=21, CMV seropositive=4). We matched participants from the first and second timepoint at which PhIP-Seq was measured with participants with scRNA-seq data. These scRNA-seq datasets were generated 5 years after the PhIP-Seq measurements, on average. In the original studies, k-nearest neighbor clustering was used to cluster the cells. Here, we performed automated cell type classification using Azimuth to annotate the cells (68). In detail, we conducted a supervised analysis guided by a reference dataset to enumerate cell types that would be challenging to define with an unsupervised approach. We mapped our scRNA-seq query dataset to a recently published CITE-seq reference dataset of 162,000 PBMCs measured with 228 antibodies (68). For this process, we used a set of specific functions from the Seurat R package v4.0.0 (68,69). First, we normalized the reference dataset using the SCTransform function. Next, we found anchors between reference and query using a precomputed supervised PCA transformation through the FindTransferAnchors function. We then transferred cell-type labels and protein data from the reference to the query and projected the query data onto the UMAP structure of the reference. For these last two steps, we used the FindTransferAnchors function. The Oelen2022 dataset, which has a larger sample size and balanced representation of CMV serostatus, was considered the primary dataset to investigate alterations in cellular composition and gene expression linked to CMV serostatus. It was also used for the CMV serostatus prediction analysis. The Wijst2018 dataset, which has a smaller participant pool and an unbalanced distribution of CMV serostatus, was used only for replication.

We used both the low (l1)- and high (l2)-resolution cell-type annotations predicted by Azimuth for the Oelen2022 dataset (n=94) to closely reflect the resolution of the measured blood cell counts and bulk RNA-seq deconvolution-predicted blood cell counts. To calculate cell-type-proportions relative to the total PBMCs, we only considered cell types with >5 cells per donor in at least 5 donors. To account for the compositional nature of the scRNA-seq data, we used the CLR transformation (61), i.e., per donor, we divided cell-type-proportions by their geometric mean. For each cell type, we then fitted a linear mixed model that controls for both biological (sex and age) and technical (10X Chromium Single Cell 3’ chemistry and experimental batch) covariates (Equation 2).

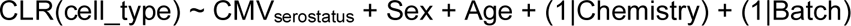

**Equation 2**. Cell type proportion model. CLR-transformed cell_type is the CLR-transformed cell-type proportion being tested. CMV_serostatus_ is the CMV serostatus prediction per participant. Chemistry is the 10X Chromium Single Cell 3’ chemistry. Batch is the experimental batch (day of the scRNA-seq library prep) data.

To test for interactions, we included either a CMV serostatus_i x Sex or a CMV serostatus_i x Age term in the model. Cell types were considered to be significantly associated with CMV seropositivity when the cellular composition change was significant at an FDR<0.05.

We then interrogated whether B3GAT1+ cells from any cell type were increased with CMV seropositivity (Equation 3). We defined B3GAT1+ and B3GAT1-subpopulations using the single-cell expression of the CD57-encoding gene (*B3GAT1*).

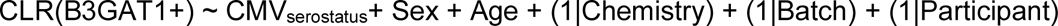

**Equation 3**. Analysis of B3GAT1+ cells. An additional random-effect term, (1|Participant), is included into Equation 2 to control for sample origin.

A second model was fitted to determine whether the CMV effect was different in different B3GAT1+ cell populations (Equation 4).

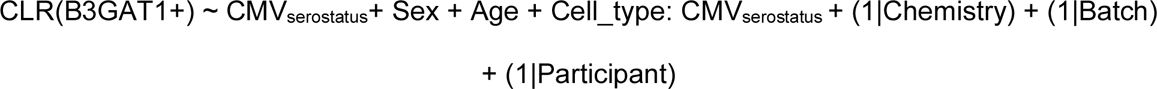

**Equation 4**. Analysis of B3GAT1+ cells. An additional fixed-effect term, Cell_type: CMV_serostatus_, is included into Equation 3 to account for CMV–cell type interactions.

The models in Equation 3 and Equation 4 were then compared using a likelihood ratio test to estimate whether the effect was different from cell type. In a subsequent analysis, we ran Equation 2 per cell type in B3GAT1+ cells separately.

#### DGE analysis with CMV serostatus

We performed pseudobulk DGE analyses between CMV seropositive and seronegative individuals from Oelen2022 (n=94) at the high (l2)-resolution cell-type annotations predicted by Azimuth. Additionally, we defined the B3GAT1+/− subpopulations using the single-cell expression of this CD57-encoding gene (*B3GAT1*). First, we aggregated (summed) the single-cell gene expression profile per donor and cell-type combination using the sparse_Sums function from the textTinyR R package v1.1.7. Only genes with a minimum expression level (>0.1 counts per million (CPM) in at least 5 donors) were tested in the pseuodbulk DGE analyses.

Next, we performed library size normalization on the pseudobulk gene expression profiles using the DGEList and calcNormFactors from the edgeR R package v3.38.4(70). Afterwards, we used the voomWithDreamWeights function from the variancePartition R package v1.28.9 to transform count data to log2-CPM (logCPM) and estimate the mean-variance relationship, which we used to compute appropriate observation-level weights. Lastly, per cell type, we fitted the following linear mixed model for CMV serostatus–DGE analysis using the dream function from the variancePartition R package v1.28.9 (Equation 5).

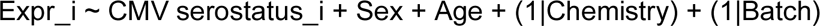

**Equation 5**. Differential expression analysis. Expr_i is the log-normalized, scaled expression of the gene being tested in donor i. CMV serostatus_i is the CMV serostatus prediction in donor i. Sex and Age are phenotypic variables from the donors. Chemistry is the 10X Chromium Single Cell 3’ chemistry. Batch is the experimental batch (day of the scRNA-seq library prep).

Genes were considered to be differentially expressed with CMV seropositivity when the gene expression change was significant at an FDR<0.05. A binomial test using the binom.test function from the stats R package v4.3.0 was used to assess whether there was a significant preference for up- or down-regulated DEGs. The Jaccard index was computed to determine sharing among the cell-type-specific DEGs (Equation 6).

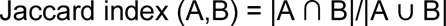

**Equation 6**. Jaccard index. A and B are the set of cell-type-specific DEGs. |A ∩ B| is the size of the intersection between A and B sets. |A ∪ B| is the size of the union between A and B sets.

Cell-type-specific up- and down-DEGs (with an absolute log-fold-change > 0.5) were used to compute an up- and down-module score using the AddModuleScore function from the Seurat R package v4.0.0 (68,69), which calculates the average expression levels of each program (up- and down-DEGs) on single-cell level, subtracted by the aggregated expression of control feature sets. Later, per each single cell type, we combined the two scores by subtracting the down-DEGs scores from the up-DEGs scores (ΔDE score).

#### Functional enrichment analyses from the CMV serostatus–DGE analysis

We performed a functional enrichment analysis through an over-representation analysis using WebGestalt (WEB-based GEne SeT AnaLysis Toolkit) (71). As the input gene list, we used the 6,262 and 1,918 DE genes identified in CD4+ CTL and CD8+ TEM cells, respectively, split by their direction of effect (i.e., positively or negatively associated with CMV seropositivity). As the background gene set, we used the 10,687 and 13,579 expressed genes that were tested in the pseudobulk DGE analyses of the CD4+ CTL and CD8+ TEM cells, respectively. As the functional database, we used the gene ontology (GO) biological process pathway database. Advanced default parameters were used (minimum and maximum number of genes for a category: 5 and 2000, multiple test adjustment: Benjamini-Hochberg, significance level: FDR ≤ 0.05, number of categories expected from set cover: 10). Finally, we used two functions from the rrvgo R package v1.12.2 to simplify the redundancy of GO sets by grouping similar terms based on their semantic similarity. The calculateSimMatrix function allowed us to get the similarity matrix between terms, and the reduceSimMatrix function grouped terms, based on the similarity matrix, with at least a similarity below a specific threshold (i.e., 0.9) and selected the term with the higher score (i.e., score reflecting how unique the term is) within the group as the group representative.

#### CMV serostatus prediction analysis

Cellular composition and gene expression profiles from Oelen_2022 (n=94) were used to train a model to predict CMV serostatus. We performed five times a 3-fold cross-validation fitting a logistic regression model with a lasso penalty using the V2 (n=64, CMV seronegative=43, CMV seropositive=21) and V3 (n=30, CMV seronegative=16, CMV seropositive=14) data independently. We used the cv.glmnet function from glmnet v4.1.8 (72) to train and test a lasso regression model (α=1). To determine the hyperparemeter λ, which represents the strength of shrinkage, a 4-fold cross-validation was run on the training data, and the lowest average mean squared error was used for hyperparameter selection. The lasso regression model with best λ was then trained and used to predict response value in the held-out data. For the cellular composition model, we used cell-type proportions as features. We only considered cell types with >5 cells per donor in at least 5 donors. For the gene expression models, we trained a model per l2-subpopulation using all expressed genes (>0.1 CPM in at least 5 donors) as features. To validate our best-performing model (the one based on cellular composition), we used CMV annotated V2 data from an independent study (Wijst2018) (n=25, CMV seronegative=21, CMV seropositive=4). In this case, we combined the V2 and V3 data from Oelen2022 (n=94, CMV seronegative=59, CMV seropositive=35) to achieve a larger and more balanced training dataset. First, we merged the cell-type-proportions from the three datasets (Oelen2022 V2, Oelen2022 V3 and Wijst2018) and extracted the residuals of Equation 7.

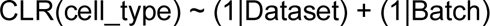

**Equation 7**. Batch correction. CLR-transformed cell type is the CLR-transformed cell type proportion being tested. Dataset is V2/V3 Oelen2022 or Wijst2018. Batch is the experimental batch (day of the scRNA-seq library prep).

Next, we split the Oelen2022 data into 80% training and 20% testing sets using the createDataPartition function from the caret R package v6.0.94. CMV serostatus was predicted among the 20% testing dataset from Oelen2022 and the independent Wijist2018 dataset.

The performance metrics (ROC AUC, F1 score, Matthews Correlation Coefficient (MCC) and accuracy) were computed using the roc_auc and metrics_set functions from the yardstick R package v1.2.0. Confusion matrices were also calculated using the same R package.

#### Rhinovirus association analysis

Per TL, we ran a linear regression with lasso penalty using all PhIP-Seq peptides annotated as of rhinoviral origin as covariates and TL as dependent variable, while performing a 5-fold cross-validation for determination of the λ parameter. This procedure was repeated 100 times. We then assessed which peptides were more often selected in each analysis to identify the best peptide for association with TL. Among the most-often-selected peptides, we ran a linear model using all TLs as dependent variables and including a covariate specifying cell type and a random effect on the intercept specifying sample ID. We then compared the fit of models including basic covariates (age, sex), CMV infection and smoking habits (current smoker and ever smoker). In addition, in a subsequent analysis, we used a binary definition of rhinovirus infection, using the top selected peptide from the lasso regression (*twist_35344*), and matched samples with and without immune response against this peptide according to their age and sex (R MatchIt v4.3.4, method ‘nearest’, 1:1 ratio).

We performed similar analyses as described for CMV association between the top rhinoviral feature and TLs, cell counts and predicted cell counts. A mediation analysis was performed using regmed v2.0.4 and mediation v4.5.0.

#### Assessment of statistical significance

For univariate association models, we used an FDR estimated by calculating the total number of positives and false positives from a null distribution of permuted P-values. For individual terms from a linear model, we permuted the term where the FDR was to be estimated. For interaction effects, we permuted both the interaction terms that make up the interaction. Positives were defined as the number of P-values ≤ a specific threshold in the distribution of non-permuted P-values. False positives were defined as the number of P-values from the null distribution ≤ a given non-permuted P-value, divided by the number of permutations performed (usually 1,000). The FDR for such a threshold was then defined as the ratio of false positives to positives.

We used an FDR_per of at least 0.05 as a significance threshold, but other trends that did not reach this threshold were also analyzed.

In the analyses of single-cell data, we used Benjamini-Hochberg FDR estimation.

## Data availability

The data presented were collected by the Lifelines cohort study. Lifelines is specifically organized to make assessment results available for (re)use by third parties.

- PhIP-Seq: Raw and processed, previously generated PhIP-Seq data are available in the European Genome-Phenome Archive (EGA) under the accession EGA: EGAS00001006999.
- Biological aging data, including sj-TRECs expression, methylation age predictions and TL measurements, in addition to cytokines and chemokine measurements, cell-type counts and predictions, can be requested through Lifelines. A research proposal must be submitted for evaluation by the Lifelines Research Office.
- Processed (de-anonymized) scRNA-seq data, including a text file that links each cell barcode to its respective individual, has been deposited at the EGA, which is hosted by the EBI and the CRG, under accession number EGAS00001005376 for Oelen2022 and EGAS00001002560 for Wijst2019.
- In addition to Lifelines data, we used data from the 1000IBD cohort study to train models for CMV prediction. PhIP-Seq data is available at EGA: EGAD00001004194.
- The 300BCG cohort was used for replication of several analyses. Telomere measurements for this cohort are available upon contact.

## Code availability

Code used for analysis can be found in the public GitHub repository: https://github.com/GRONINGEN-MICROBIOME-CENTRE/Phip-Seq_LLD-IBD/tree/main/ImmunoSenescence

## Author contribution

S.A-S designed the project, performed the analyses, supervised the analyses carried out by other team members and wrote the original manuscript.

A.R-C designed and performed the single-cell analysis and wrote the original manuscript, M.G.P.W, M.J. and M.M supervised and designed single-cell analyses.

L.F. funded the single cell experiments and analysis.

A.C. partially performed CMV analyses and wrote the draft of the original manuscript.

A.R.B. provided feedback and interpretation to biological findings

O.B. and M.N. provided replication in 3000BCG.

G.A. and P.L. provided feedback regarding TL measurements and biology in relation to CMV infection.

T.V. provided feedback and developed ideas regarding PhIP-Seq analyses.

D.V.B. provided feedback on the epidemiological implications of CMV and other pathogens.

J.F. and A.Z. provided overall feedback and helped in project design.

All authors read and agreed on the contents of this manuscript.

## Acknowledgments

We thank K. Mc Intyre for English editing. The Lifelines Biobank initiative has been made possible by a subsidy from the Dutch Ministry of Health, Welfare and Sport; the Dutch Ministry of Economic Affairs; the University Medical Center Groningen; the University of Groningen; and the Northern Provinces of the Netherlands. The authors wish to acknowledge the services of the Lifelines Cohort Study, the contributing research centers delivering data to Lifelines, and all the study participants.

## Funding information

A.Z.: Netherlands Heart Foundation (IN-CONTROL CVON 2018-27). EU Horizon Europe Program grant INITIALISE (101094099). NWO Gravitation project ExposomeNL (024.004.017) and NWO-VIDI (016.178.056).

J.F.: Netherlands Heart Foundation (IN-CONTROL CVON 2012−03). NWO-VIDI (864.13.013). NWO-VICI (VI.C.202.022). ERC Consolidator Grant (grant agreement no. 101001678).

T.V.: European Union - ERC STG, project number 101075733.

M.M.: RYC-2017-22249. PID2019-107937GA-I00.

A.R.: FPI [grant no. PRE2019−090193].

M.W.: Dutch Research Council (NWO-VENI 192.029 and NWO-VIDI 223.041)

A.R.B.: NWO Rubicon grant [grant no. 452022317].

## Supplementary Material

### Supplementary Results

**Supplementary Figure 1.**
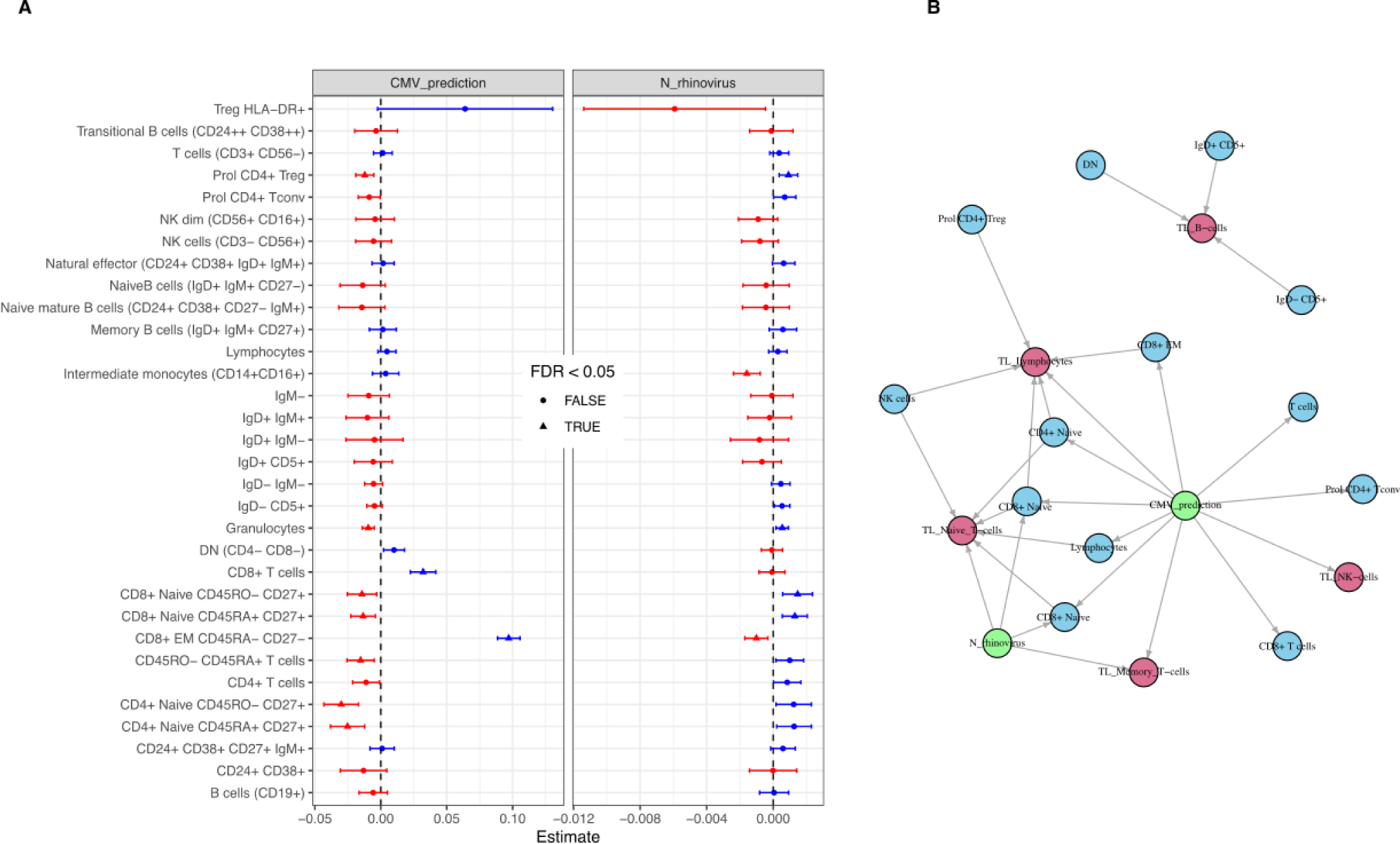
Rhinovirus and CMV signal on cell composition. **A.** Estimated effect size of CMV prediction and breadth of rhinovirus peptides in deconvoluted cell populations (ALR-normalized), controlled for age and sex. Color indicates the direction of the effect (red: negative, blue: positive). Triangles indicate FDR_BH_<0.05 associations. Circles indicate an FDR_100perm_≥0.05. Error bars represent 95% confidence interval from the estimate. **B.** Mediation network built using Regmed. Green nodes show exposures, CMV prediction and breadth of rhinoviral antibodies (N_rhinovirus). Edges bind exposures with 1. mediators (blue), representing deconvoluted cell proportions, and 2. outcomes (red), representing TLs. Mediation effects are visualized as edges binding exposures with mediators and mediators with outcomes.

**Supplementary Figure 2.**
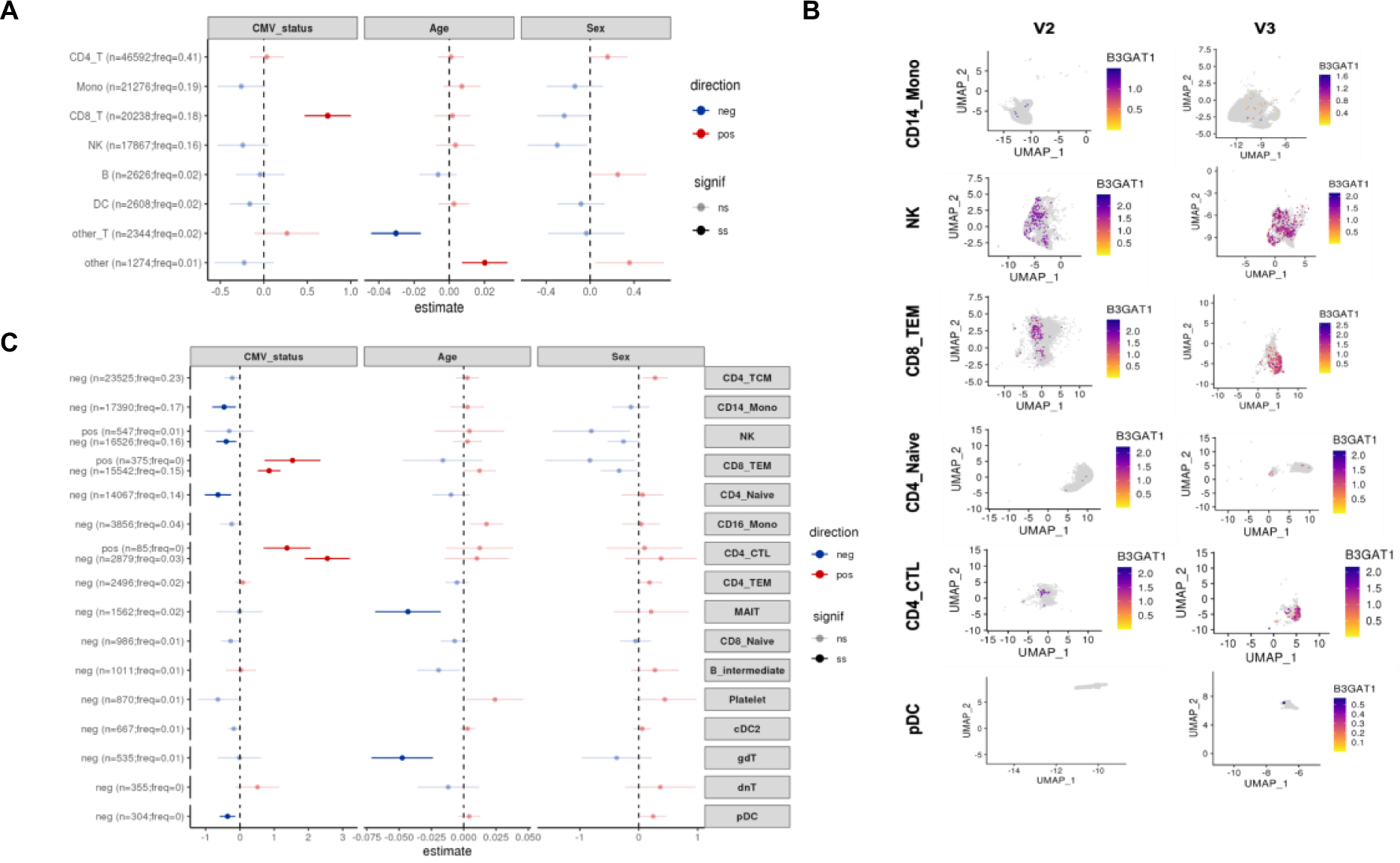
Cellular composition changes associated to CMV seropositivity. **A.** Forest plot showing the linear association between CMV serostatus, age or sex, and low-level (Azimuth’s l1) cell-type-proportions from the Oelen2022 scRNA-seq data. Estimated effect values accounting for both biological (sex and age) and technical (10X Chromium Single Cell 3’ chemistry and experimental batch) covariates are displayed. Error bars represent the 95% confidence interval of the estimated effect. Color indicates the sign of the estimated effect values (red: positive, blue: negative). Shading represents the statistical significance of the estimated effect values (darker: significant, FDR≤0.05; lighter: non-significant, FDR>0.5). The absolute number and relative frequency of each cell type is shown. **B.** UMAPs showing the expression of *B3GAT1* (CD57-encoding gene) in each of the high-level (Azimuth’s l2) cell types altered by CMV seropositivity in (C). **C.** Forest plot showing the linear association between CMV serostatus, age or sex, and high-level (Azimuth’s l2) cell-type-proportions from the Oelen2022 scRNA-seq data, distinguishing *B3GAT1*+/− populations. Estimated effect values accounting for both biological (sex and age) and technical (10X Chromium Single Cell 3’ chemistry and experimental batch) covariates are displayed. Error bars represent the 95% confidence interval of the estimated effect. Color indicates the sign of the estimated effect values (red: positive, blue: negative). Shading indicates the statistical significance of the estimated effect values (darker: significant, FDR≤0.05; lighter: non-significant, FDR>0.5). The absolute number and relative frequency of each cell type is shown.

**Supplementary Figure 3.**
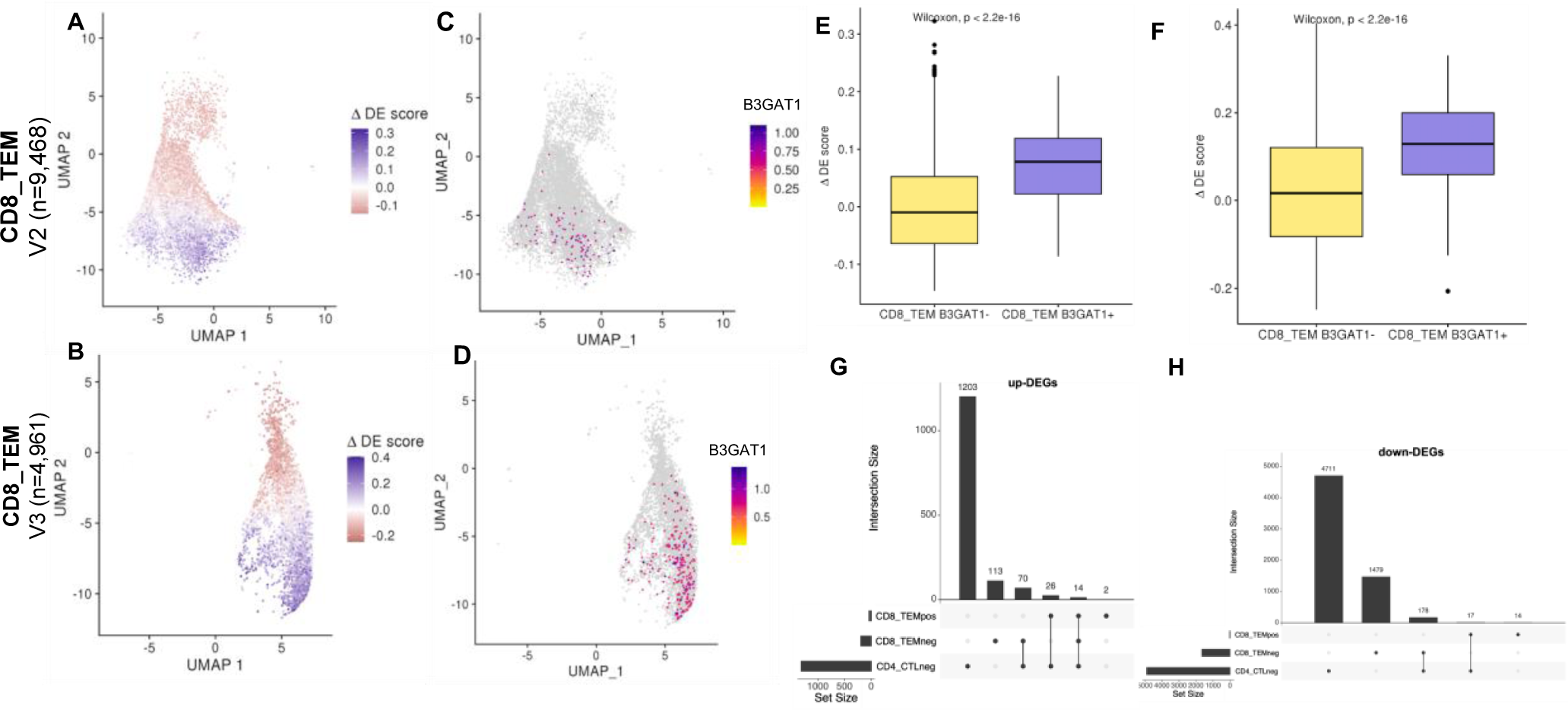
**A–B** UMAPs showing the ΔDE (differential expression) scores using the Oelen2022 scRNA-seq dataset: CD8+ TEM cells in V2 (**A**) and V3 (**B**) data. **C–D**. UMAPs showing the expression of *B3GAT1* (CD57-encoding gene) in CD8+ TEM cells using the Oelen2022 scRNA-seq V2 (**C**) and V3 (**D**) data. **E–F.** Boxplots showing the differences between the distribution of ΔDE scores in CD8+ TEM B3GAT1+ and CD8+ TEM B3GAT1-cells using the Oelen2022 scRNA-seq V2 (**E**) and V3 (**F**) data. **G–H.** Upset plots showing the number of up-regulated (**G**) and down-regulated (**H**) DEGs shared or unique among the high-level (Azimuth’s l2) B3GAT1+/− cell types using the Oelen 2022 scRNA-seq data. ‘pos’ and ‘neg’ after the l2 cell-type refer to ‘B3GAT1+’ and ‘B3GAT1‘-’, respectively.

**Supplementary Figure 4.**
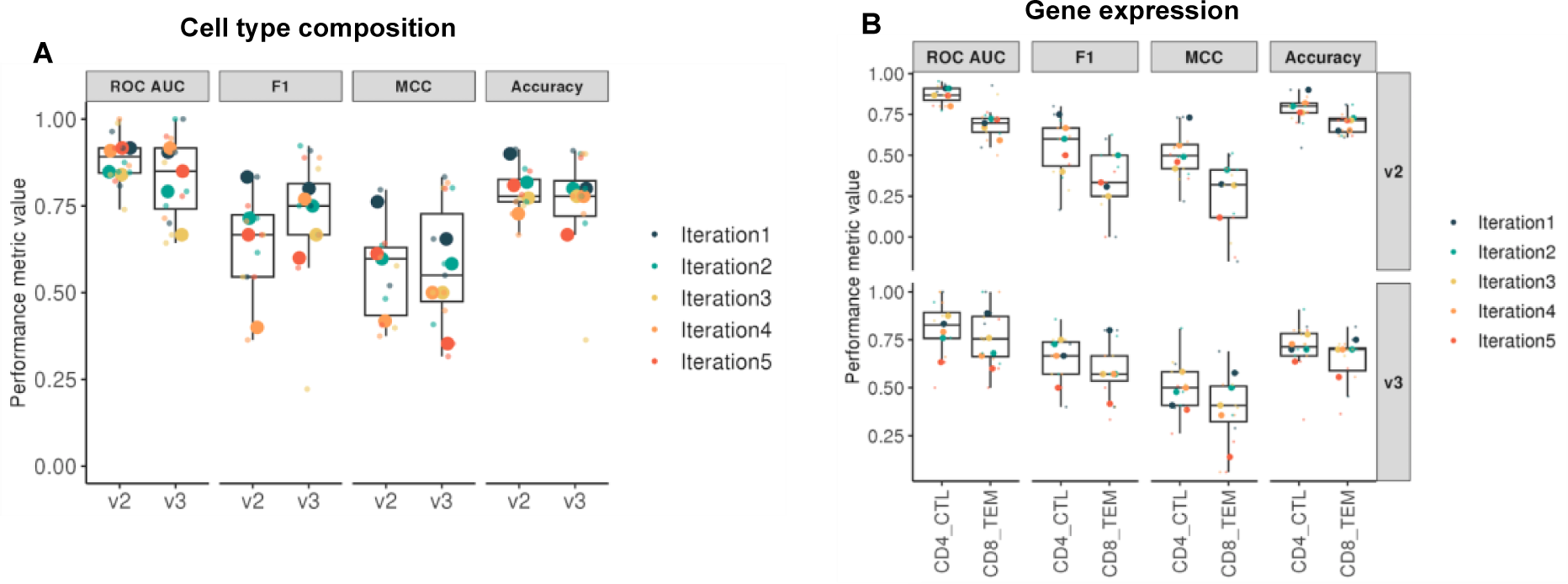
Boxplots showing the performance of the CMV serostatus prediction model (using Oelen2022 V2 or V3 data) based on cellular composition (**A**) or the gene expression profile of CD4+ CTL or CD8+ TEM cells (**B**). We performed five times (iterations) a three-fold cross-validation fitting a logistic regression model with a lasso penalty. Color indicates the different iterations. Small symbols correspond to each of the cross-validation folds. Larger symbols correspond to the median across the three cross-validation folds in each iteration. Performance metrics used were: ROC AUC, F1 score, Matthews Correlation Coefficient (MCC) and accuracy.

