## Supplementary Material for "Antibody signatures against viruses and microbiome reflect past and chronic exposures and associate with aging and inflammation"

### Results

**Antibody responses against rhinoviruses are correlated with longer telomeres and cellular composition changes**

We explored the observed association of antibodies against rhinovirus and longer telomeres. Due to the lack of a clear selection criterion for a single representative rhinoviral peptide, we employed a regularized linear model (lasso penalty) regression on different bootstraps of the complete dataset to identify rhinoviral peptides that were consistently associated with different telomere lengths (TLs). Our analysis revealed that one individual peptide, *twist_35344*, targeting ‘polyprotein & VP1 capsid protein’ and a score representing the breadth of antibody-bound peptides of rhinoviral origin, total number of anti-rhinovirus antibodies, were frequently selected as features in the regularized model (median among cell types of 93% and 75.8%, respectively). We then compared TLs from all individuals with the breadth of antibody-bound rhinoviral proteins and *twist_35344* and found that TLs were more strongly associated with the breadth of antibody-bound rhinoviral proteins than with *twist_35344* (linear-mixed model of the effect on all TL, P_breadth_antibody_=1.8x10^-11^, P_twist_35344_=1.38x10^-5^). Consequently, we utilized the count score representing the breadth of rhinoviral antibodies to investigate the associations with TL.

It is worth noting that rhinoviral antibody-bound peptides are often observed in younger individuals (1,2), while cytomegalovirus (CMV) infections are more prevalent at older age. To address the potential confounding effect of age, we adjusted for CMV status and compared the strength of the association with and without adjustment for age (which was treated as a categorical variable to account for possible non-linear effects). Remarkably, the association between the breadth of rhinoviral antibodies and TL remained significant after adjusting for age (linear-mixed model of the effect on all TL, average effect breadth rhinovirus antibodies in all TLs without accounting for age=2.688x10^-2^, P=5.82x10^-10^; average effect breadth rhinovirus antibodies in all TLs accounting for age effect=1.677x10^-2^, P=2.70x10^-5^), indicating that the association of the presence of rhinoviral antibodies to longer telomeres is partially independent of participant age. Similarly, after matching 393 participants with antibody responses against *twist_35344* (rhinoviral ‘polyprotein & VP1 capsid protein’) with those with no antibody response, based on the nearest age-sex match (see **Methods**), we still identified significant positive effects of rhinovirus on TLs (multivariable model with age, CMV and *twist_35344,* effect_matched_=0.21, P_matched_=2.5x10^-4^, effect_notmatched_=0.173, P_notmatched_=4.27x10^-4^). Furthermore, we investigated the independence of the rhinoviral association from smoking, as smoking is often associated with increased rhinoviral infections and typically considered a factor negatively associated with TL. Our analysis revealed that the association between the breadth of rhinoviral antibodies and TL remained significant after adjusting for smoking (effect on all TL, effect=1.841x10^-2^, P=4.61x10^-6^).

Overall, after accounting for CMV, smoking habits, age and sex, the breadth of rhinoviral antibodies was associated with all TLs (P<1.5x10^-03^), but this effect was significantly different between cell types (likelihood ratio test (LRT) model with interaction term of cell type and rhinovirus vs model without, P=8.576x10^-7^). With respect to TLs, rhinoviruses were more strongly associated with the TLs of memory T-cells (estimate=0.02, P=4.05x10^-6^), lymphocytes (estimate=0.019, P=3x10^-5^) and naïve T-cells (estimate=0.019, P=4.61 x10^-5^) [**Sup Table 4**]. We did not find statistical significance supporting differences between age groups (LRT model with age group interaction with rhinovirus vs model without, P=0.46).

We also observed that the breadth of rhinoviral antibody responses was significantly associated with specific cell populations, mirroring those attributed to CMV, but in the opposite direction [**Sup. Table 4**]. For example, we found significant associations between rhinoviral antibody breadth and intermediate monocytes (CD14+CD16+) (effect=-1.6x10^-3^, P=1.1x10^-4^), CD8+ naïve cells (effect=1.29x10^-3^, P=7.4x10^-4^) and proliferative CD4+ Treg cells (effect=9.1x10^-4^, P=1.3x10^-3^), among others [**Sup. Fig 1A**]. To explore the mediating role of cell composition and TL in these associations, we conducted a mediation analysis that included both CMV infection and the breadth of rhinovirus antibodies. The results suggest that CMV and rhinoviral effects are independent and that cell composition partially mediated the changes in TL [**Sup. Fig 1B**]. Specifically, the effect of rhinovirus on TL in naïve T-cells was found to be partially mediated by the predicted cell counts of CD8+ naïve cells, accounting for 17.1% (95% CI, 0.07–0.35) of its effect on TL.

**Cell composition association to CMV serostatus in single-cell data**

In our previous results using measured cell counts and cell counts predicted from bulk RNA-seq, we found a CMV-associated expansion of CD8+ T-cells, particularly CD8+ EM, and a decrease of proliferative and naïve CD4+ T-cells. Using scRNA-seq data, we used both the low (l1)- and high (l2)-resolution cell-type-annotations predicted by Azimuth (3) to classify cells in order to closely reflect the resolution of the measured and deconvoluted blood cell counts (see **Methods**). At l1 level, we replicated the previously observed association [**Fig 3B**] between CD8+ T-cells and CMV serostatus (effect=0.74, P=4x10^-7^, FDR=3.2x10^-6^) [**Sup. Fig 2A**]. At l2 level, we identified four significant associations (FDR<0.05) [**Fig 4A**] and replicated three previously observed cell proportion–CMV associations [**Fig 3B**]: the negative association of CMV with CD4+ naïve T-cells (effect=-0.56, P=3.7x10^-3^, FDR=2.2x10^-2^), which were previously reported to be reduced by CMV (4), a positive association between CMV and CD8+ effector memory T-cells (TEM) (effect=0.95, P=32.3x10^-6^, FDR=2.7x10^-5^), a subpopulation able to expand and generate TEFF cells upon rechallenge (5); and a significant decrease of regulatory T-cells (Treg) (effect=-0.52, P=2.2x10^-3^, FDR=1.8x10^-2^), with such a reduction previously observed in CMV-positive males (6). We found an additional association that was not seen in the deconvoluted data: an increase of CD4+ cytotoxic T lymphocytes (CTL) (effect=2.66, P=8.5x10^-12^, FDR=2.1x10^-10^) with CMV seropositivity, with CTL known to be mediators of antiviral defense (7) [**Sup. Table 5A**].

**CMV infection linked with depletion and overexpression of transcriptional pathways in CD4+ CTL and CD8+ TEM cells**

We assessed whether our reported DEGs belonged to similar functional pathways, thereby highlighting the biological interplay between CMV seropositivity and gene expression [**Fig 4E**]. To explore this, we performed a functional enrichment analysis separately for the up- and down-DEGs in each of the two cell subtypes [**Sup. Table 5D**]. Within the genes positively associated with CMV serostatus in the CD4+ CTL cells, we found an enrichment of the pathways *negative regulation of metabolic* (GO:0009892, enrichment ratio=1.55, P=2.2x10^-16^, FDR=2.2x10^-16^) and *negative regulation of gene expression* (GO:0010629, enrichment ratio=1.75, P=2.2x10^-16^, FDR=2.2x10^-16^). In addition, we found an enrichment for *translation* (GO:0006412, enrichment ratio=2.34, P=2.2x10^-16^, FDR=2.2x10^-16^) and *peptide biosynthetic process* (GO:0043043, enrichment ratio=2.3, P=2.2x10^-16^, FDR=2.2x10^-16^). Unlike the many viruses that suppress cellular protein synthesis, CMV stimulates host mRNA translation and polyribosome formation, even in uninfected cells (8). Conversely, we found a down-regulation of several lipid biosynthetic processes, including *lipid biosynthetic process* (GO:0008610, enrichment ratio=1.27, P=1.2x10^-6^, FDR=3.6x10^-3^), *phospholipid biosynthetic process* (GO:0008654, enrichment ratio=1.35, P=1.4x10^-5^, FDR=2x10^-2^) and *glycerolipid biosynthetic* *process* (GO:0045017, enrichment ratio=1.36, P=2.7x10^-5^, FDR=2x10^-2^). Systemic metabolic sequelae such as insulin resistance and dyslipidemia represent long-term health consequences of many infections (e.g., human immunodeficiency virus, hepatitis C virus and SARS-CoV-2) (9). In addition, the *anion transport* pathway (GO:0006820, enrichment ratio=1.3, P=2.7x10^-5^, FDR=2x10^-2^), which can be modulated by viral proteins (10), was negatively associated with CMV seropositivity. On the other hand, focusing on CD8+ TEM cells, many immune-related pathways, such as *regulation of leukocyte activation* (GO:0002694, enrichment ratio=3.21, P=2.2x10^-6^, FDR=1.9x10^-3^) and *immune effector process* (GO:0002252, enrichment ratio=2.57, P=1.7x10^-8^, FDR=1.3x10^-4^), were positively associated with CMV seropositivity, together with *exocytosis* (GO:0006887, enrichment ratio=2.52, P=3.3x10^-6^, FDR=2.5x10^-3^), one of the major mechanisms of cytotoxicity involved in the clearance of virus-infected cells (11). In addition, our set of negatively enriched pathways revealed a regulation of signaling receptor activity, specifically the *G protein-coupled receptor signaling* pathway (GO:0007186, enrichment ratio=1.97, P=2.9x10^-8^, FDR=1.1x10^-4^). CMV, as a member of the *Herpesviridae* family, encodes G protein-coupled receptors (GPCRs) showing homology to human chemokine receptors, which might be used as decoy receptors to prevent cytokine action (12). By means of these constitutive GPCRs, herpesviruses have devised strategies to rewire host cell-signaling pathways, thereby promoting viral biology and subsequent pathogenic effects (13). Lastly, since most of the pathways were subpopulation-specific, we explored the enriched pathways among the DEGs shared between CD4+ CTL and CD8+ TEM cells. Besides the positive association of leukocyte activation and immune effector processes with CMV seropositivity, we found the *homotypic cell-cell adhesion* pathway (GO:0034109, enrichment ratio=14.63, P=2.1x10^-5^, FDR=2.1x10^-2^) to be enriched in our set of shared DEGs. Indeed, an up-regulation of adhesion molecules has been reported to occur on activated T-cells by culture with the CMV antigen, mainly on CD45RO+ T memory cells, which may have a role in immune reaction or inflammatory modulation (14).

**CMV prediction using single-cell data**

Beyond CMV serostatus, sex and age are other major individual-specific characteristics that independently impact immune function. While previous studies have highlighted changes in cell proportions related to age, including expansions of CD16+ monocytes (15) and CD8+ TEM (16,17) and a decrease of mucosal-associated invariant T-cells (MAIT) cells (17), such studies have failed to account for the changes produced by CMV, which increases in prevalence at older ages. We therefore aimed to distinguish the contribution of each of these factors to altering circulating immune cell-type composition independently. While we found no associations with sex, we observed distinct cell types to be altered with age compared to those altered by CMV serostatus [**Fig 4A**] [**Sup. Fig 2A**]. We identified positive associations of age with the relative amount of CD16+ monocytes (effect=0.02, P=9x10^-4^, FDR=7.2x10^-3^) and platelets (effect=0.03, P=8.3x10^-3^, FDR=4x10^-2^) and negative associations with age for CD8+ naïve T-cells (effect=-0.04, P=8.4x10^-6^, FDR=2.1x10^-4^), MAIT cells (effect=-0.04, P=2x10^-3^, FDR=1.3x10^-2^) and γδ T-cells (gdT) (effect=-0.04, P=3x10^-4^, FDR=4x10^-3^). We did not find any cellular composition changes that were associated with the interaction between sex or age and CMV serostatus.

Our results indicate that CMV serostatus is an important factor that alters cell-type fractions and the transcriptome of PBMCs. Given its relation with both age and sex, CMV serostatus might act as a confounding factor in cell expression studies (if unaccounted for). Since most studies do not directly measure CMV serostatus in their participants, we assessed whether prediction models using cellular composition or gene expression profiles might be widely applied to predict CMV serostatus. We performed five times a three-fold cross-validation fitting a logistic regression model with a lasso penalty using the Oelen2022 V2 (n=64, CMV seronegative=43, CMV seropositive=21) and V3 (n=30, CMV seronegative=16, CMV seropositive=14) data (see **Methods**). We attained comparably high performances using the model trained with cell-type proportions (median AUC: V2=0.91, V3= 0.85; median F1: V2=0.7, V3=0.75) [**Sup. Fig 4A**] and the one trained with the CD4+ CTL gene expression profile (median AUC: V2=0.87, V3=0.79; median F1: V2=0.6, V3=0.7), followed by the CD8+ TEM model (median AUC: V2=0.7, V3=0.68; median F1: V2=0.32, V3=0.57) [**Sup. Fig 4B**]**.** Low performance metric scores were obtained using models trained with the gene expression of other cell subpopulations. Interestingly, CD4+ CTL was also found among the most highly predicted features in the cell-type composition model. Although all the expressed genes were used as features in the gene-expression-based models, in the CD4+ CTL model, both the magnitude of change (ρ=-0.53, P=6.2x10^-4^) and the level of significance (ρ=0.33, P=4.8x10^-4^) in the DGE analysis decided the importance of genes in the prediction model. *TNFRSF10A* and *EP400NL* were the CMV seropositivity DEGs with the highest feature importance (feature importance≥1), and they also showed a large magnitude of gene expression change (logFC=-1.24 and logFC=-1.03, respectively). *TNFRSF10A* (TRAIL receptor 1), a member of the TNF-receptor superfamily, is involved in transducing cell death signal and cell apoptosis and has been shown to be downregulated by CMV to evade extrinsic pro-apoptotic pathways (18). *EP400NL* (EP400 N-terminal-Like gene) is a pseudogene of the *EP400* gene, one of the core components of the Tip60 acetyltransferase complex that putatively interacts with CMV’s *UL27* gene (19). To validate our best-performing model (based on cell-type composition), we used CMV-annotated V2 data from an independent study, Wijst2018 (20) (n=25, CMV seronegative=21, CMV seropositive=4) (see **Methods**)**.** In this case, to acquire a more balanced and larger training dataset, we used the combined V2 and V3 data from Oelen2022 (n=94, CMV seronegative=59, CMV seropositive=35) to predict the CMV serostatus of the donors in the Wijst2018 dataset (median AUC=0.95, median F1=0.67).
