## Supplementary figures and images for "Antibody signatures against viruses and microbiome reflect past and chronic exposures and associate with aging and inflammation"

### Supplementary Fig1

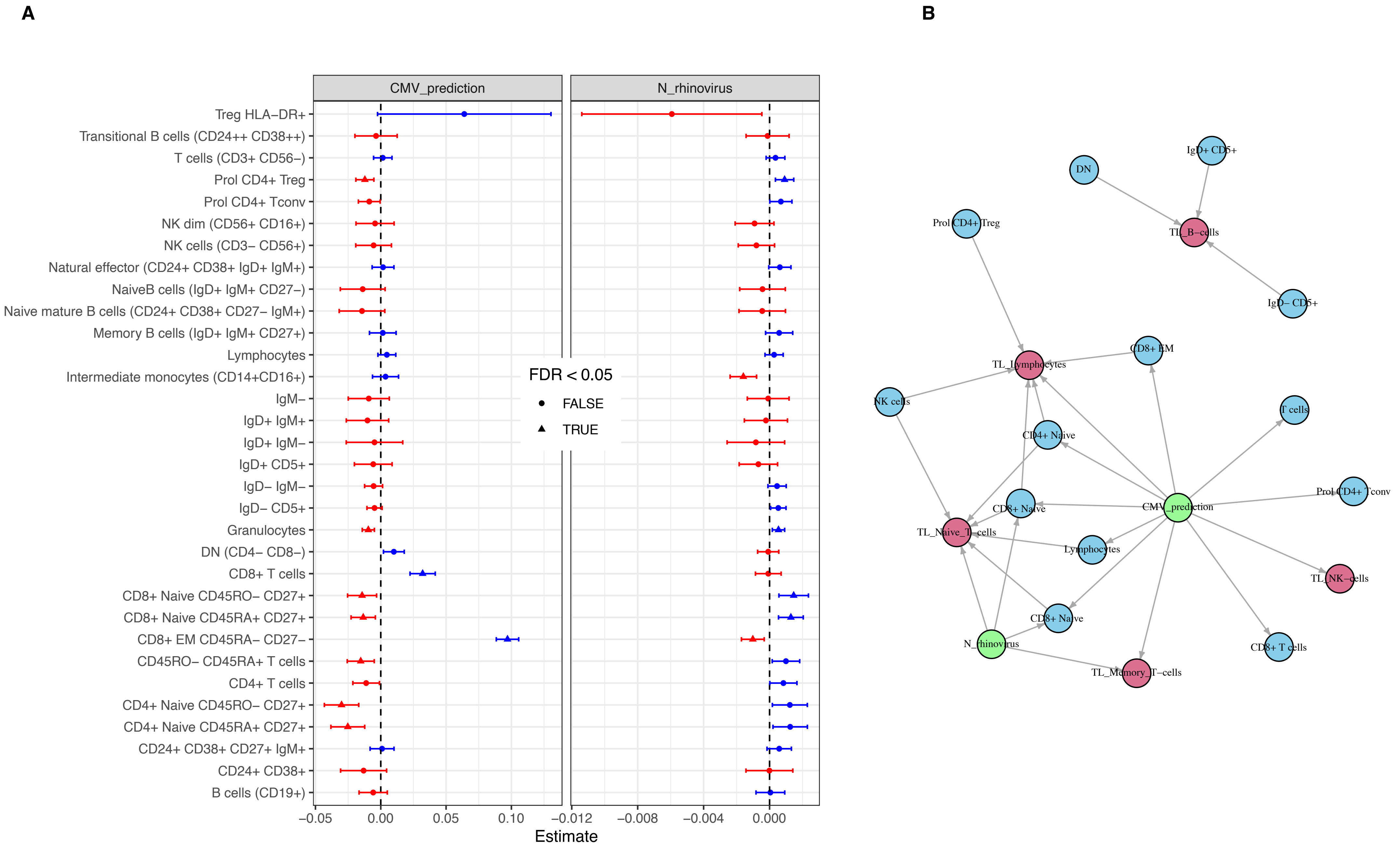

### Supplementary Fig2

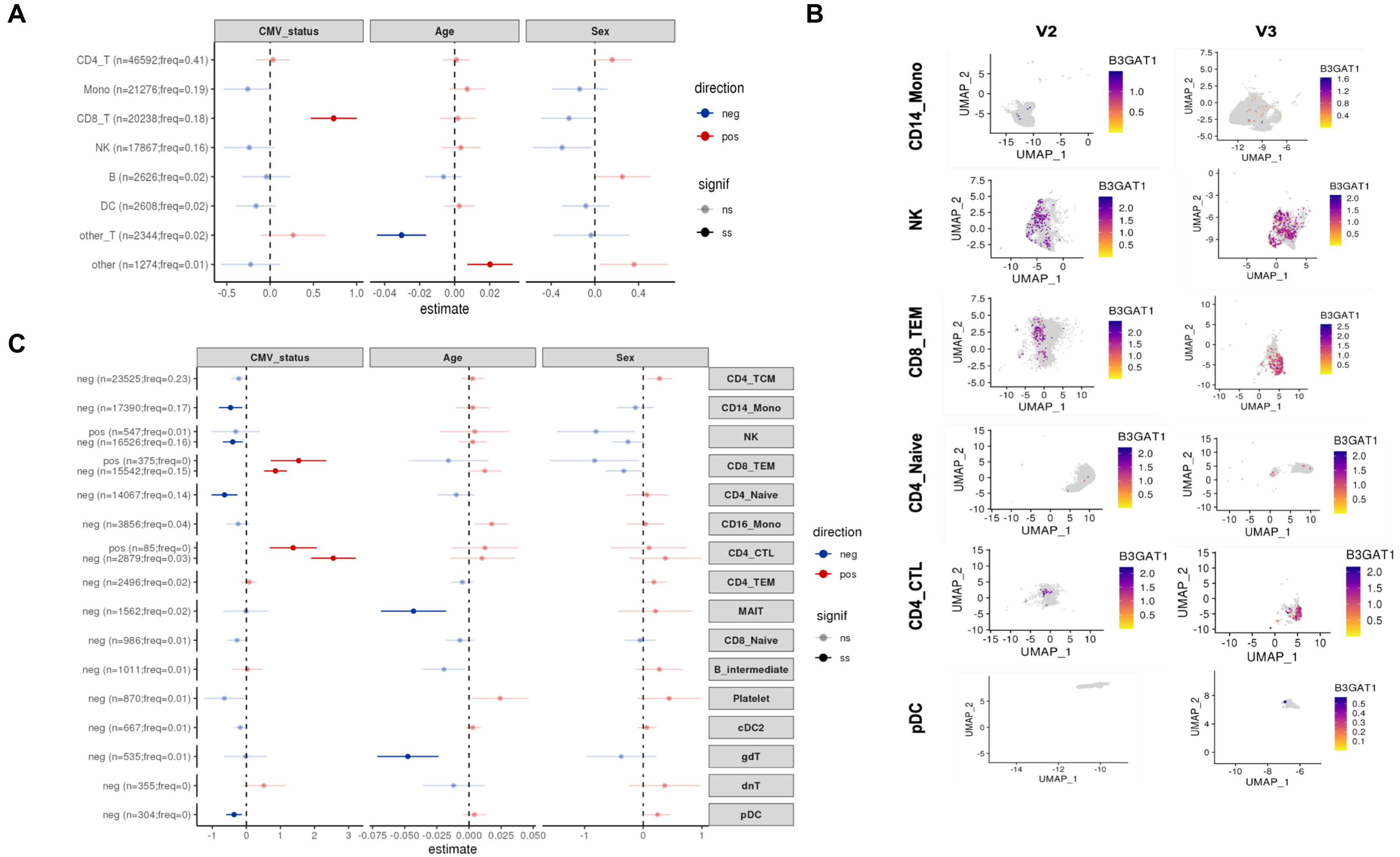

### Supplementary Fig3

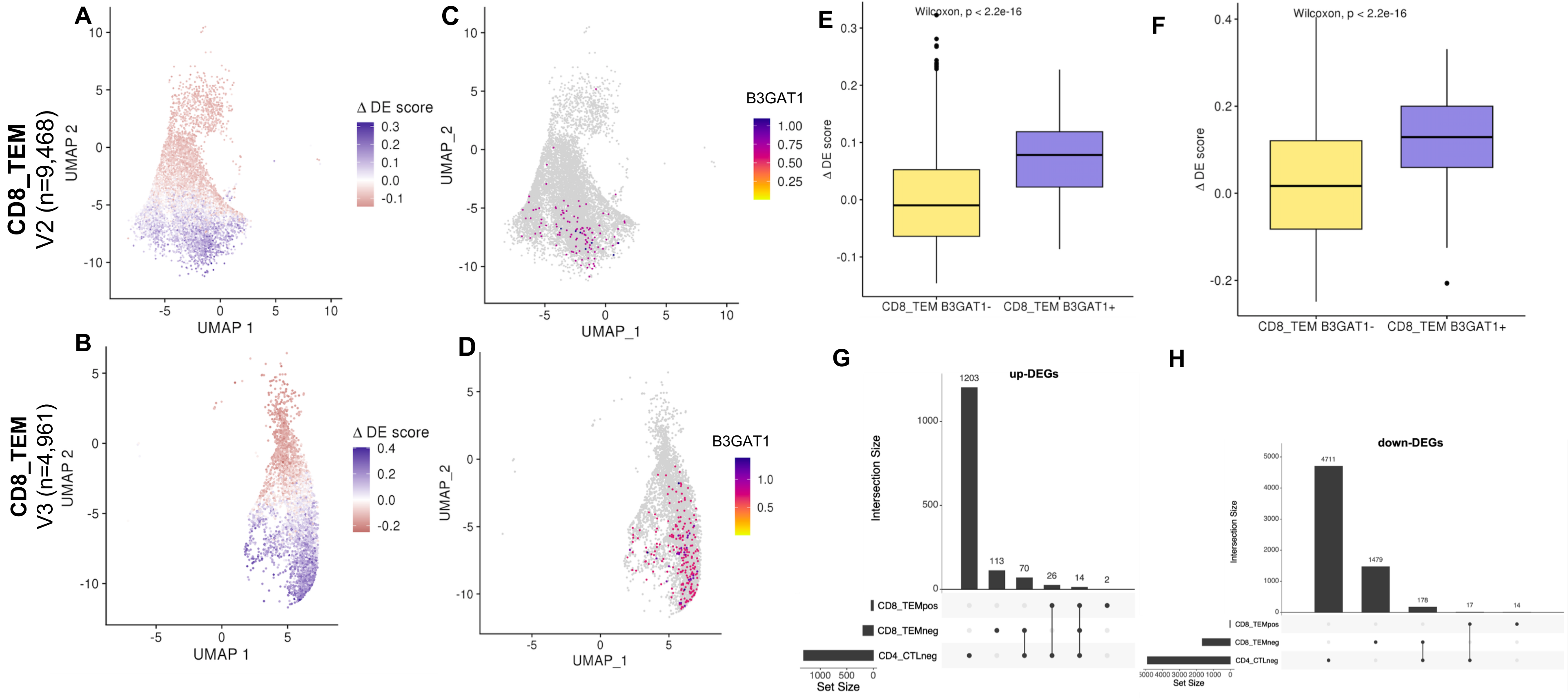

### Supplementary Fig4

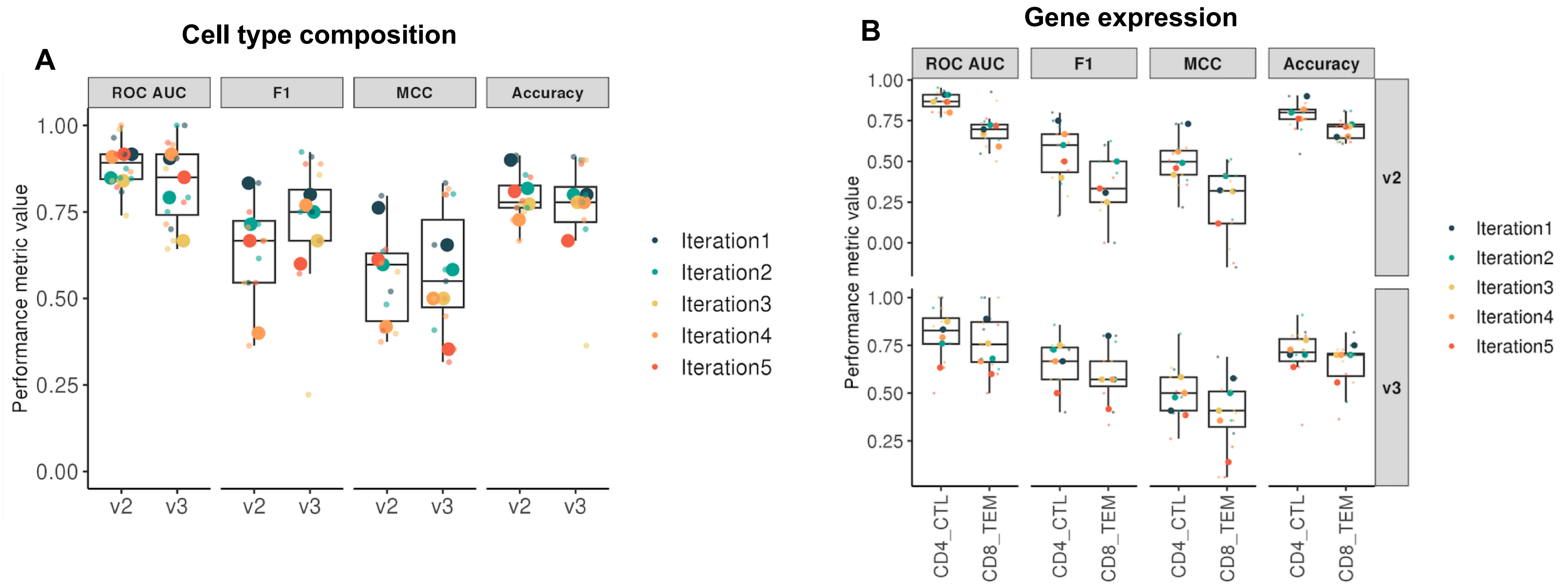
